# Proteomic and clinical biomarkers for acute mountain sickness diagnosis, prognosis, protection, and pathogenesis in a longitudinal cohort

**DOI:** 10.1101/2021.10.18.21265176

**Authors:** Jing Yang, Zhilong Jia, Xinyu Song, Jinlong Shi, Xiaojing Zhao, Kunlun He

## Abstract

Acute mountain sickness (AMS) is frequently experienced by non-high-altitude natives at high altitudes, which affects the quality of health and productivity of humans. The diagnosis of the disease mainly depends on a self-questionnaire, which reveals our insufficient understanding of AMS and the necessity of developing reliable biomarkers for AMS. In addition to 65 clinical indexes and 22 AMS symptom phenotypes, we profiled the plasma proteomic profiles of AMS via a combination of proximity extension assay with multiple reaction monitoring for a longitudinal cohort of 53 individuals divided into discovery and validation stages. Through differential analysis, machine learning models with high accuracy and protein-symptom-clinical index functional network analysis, we identified proteomic and clinical biomarkers for AMS diagnosis, prognosis, protection, and pathogenesis. RET, a top-weighted protein in the pathogenesis model, showed opposite regulations between individuals with AMS and those without AMS ascending to a high altitude. The downregulation of ADAM15 may play a protective role at high altitude in individuals without AMS. These results suggest that RET and ADAM15 could be promising therapeutic targets for AMS. Moreover, PHGDH and TRAF2 could be candidate predictive and diagnostic biomarkers for AMS, respectively. Additionally, C-peptide was found to be actively involved in the pathogenesis and could aid the assistant diagnosis of AMS. Notably, individuals with AMS showed higher gluconeogenesis activity at the plain than those without AMS. Our findings shed light on the proteomic and clinical biomarkers of AMS, provide a wealth of biological insights into AMS, and thereby promote precision medicine for AMS.

## Introduction

Acute mountain sickness (AMS), which is one of the most common types of high-altitude illness, occurs when individuals living on the plain ascend to high altitudes (above 2,500 meters). The typical symptoms of AMS are headache, gastrointestinal symptoms (poor appetite, nausea, and/or vomiting), dizziness, and fatigue. In general, the diagnosis of AMS depends on questionnaire-based diagnostic instruments, among which the Louise Lake Acute Mountain Sickness Score (LLS) is widely used(Roach et al. 2018). After exposure to the Qinghai-Tibetan Plateau, the incidence of AMS among non-high-altitude natives ranges from 31% to 57.2%(Ren et al. 2010; T.-Y. Wu et al. 2012; R. Chen et al. 2021; Gonggalanzi et al. 2017; T. Y. Wu et al. 2010). Without effective treatment, AMS may progress to life-threatening high-altitude cerebral edema or high-altitude pulmonary edema(Luks, Swenson, and Bärtsch 2017). However, the accurate diagnosis of AMS remains unclear due to an inadequate understanding of AMS(Meier et al. 2017).

The pathogenesis of AMS remains unclear and under debate. The pathophysiological mechanism of AMS involves several hypotheses, such as the induction of vasogenic and/or intracellular cerebral edema(Schoonman et al. 2008; Colleen Glyde Julian et al. 2011; S.-J. Chen et al. 2014), increases in vascular permeability due to higher levels of oxidative stress(Bailey et al. 2009), inflammation(Colleen Glyde Julian et al. 2011), increases in the vascular endothelial growth factor (VEGF) level induced by hypoxia-inducible transcription factor(Tsai et al. 2019), and a metabolic demand(Lu et al. 2018; Capitanio et al. 2017). Because these pathophysiological mechanisms cannot completely elucidate AMS, the identification of biomarkers according to these mechanisms is difficult.

The proteome is suitable for exploring the pathogenesis and discovering biomarkers of AMS. Previous studies on the proteome of AMS mainly used gel electrophoresis and mass spectrometry (MS) technology(Sharma, Sethy, and Bhargava 2013; Colleen G. Julian et al. 2014; Tyagi et al. 2014; Padhy et al. 2017; Paul et al. 2018; Jain, Ahmad, and Bhargava 2018). Notably, the gel electrophoresis-based method has several drawbacks, such as quantitative reproducibility and a biased proteome profile(Abdallah et al. 2012). Julian et al. found that the abundance of antioxidant proteins was higher in patients with AMS but not in individuals with resistance to AMS using a 20 volunteer cohort(Colleen G. Julian et al. 2014). Lu et al. revealed that proteins related to the tricarboxylic acid cycle (for example, PDHA1, SDHA1, and SUCLG1), glycolysis (such as FBP1, ALDOA, and PGK1), the ribosome, and the proteasome (for example, PSMC3) were significantly suppressed in the AMS-resistant group compared with their levels in the AMS-susceptible group via isobaric tags for relative and absolute quantitation MS(Lu et al. 2018). The levels of anti-inflammatory and/or anti-permeability factors, such as IL-1RA, HSP-70, and adrenomedullin, are higher in AMS-resistant subjects than in AMS-susceptible subjects, whereas the levels of chemotactic factors, CCL2 and TNF-α, are independent of the AMS status(Colleen Glyde Julian et al. 2011). However, the findings obtained in most of these studies were not validated. Padhy et al. observed a lower abundance of angiotensinogen and angiotensin II in high-altitude natives by MALDI_TOF/TOF(Padhy et al. 2017).

Moreover, studying the proteome of high-altitude natives could also shed light on AMS in non-high-altitude natives. Du et al. performed a TMT-label-based plasma proteomic analysis and found that CCL18, C9, and S100A9 were upregulated and that HRG and F11 were downregulated in high-altitude natives compared with non-high-altitude natives at high altitude(Du et al. 2019). Accordingly, a systematic study of the pathogenesis, candidate therapeutic targets, and protective, predictive and diagnostic biomarkers of AMS using a larger cohort and new advanced proteome technology is needed and valuable.

Proximity extension assay (PEA) technology uses antigen-antibody binding and qPCR technology to perform qualitative and quantitative analysis of proteins and has been widely used for the study of several diseases, such as COVID-19(Del Valle et al. 2020; Consiglio et al. 2020) and cardiovascular disease(Hoogeveen et al. 2020; Wallentin et al. 2021). The Olink panels cover biomarkers of crucial diseases and proteins of important biological processes involving different systems, which allows us to profile over thousands of proteins using only a small volume of plasma. Moreover, multiple reaction monitoring (MRM) is an MS-based method targeting selective peptides for protein detection and quantitation with good reproducibility and sensitivity. MRM technology is suitable for the validation of candidate biomarkers. The combination of antibody-based PEA and MS-based MRM technologies will largely eliminate false-negative signals. Therefore, we applied both PEA and MRM technologies in our study to obtain a reliable and robust result.

In this study, we systematically explored AMS using the proteomes, 65 clinical indexes, and 20 AMS symptom phenotypes of 106 plasma samples from a Chinese Han cohort consisting of 53 participants (**Fig. 1A**). We characterized the protein profile of 10 participants with AMS at plain and high altitudes using Olink’s PEA technology (**Fig. 1B**) and validated 102 key proteins using MRM proteomics technology with expanded samples and groups (**Fig. 1C**). Moreover, we identified candidate pathogenesis-related proteins and protective, predictive, and diagnostic biomarkers of AMS via statistical analysis and a machine learning-based model (**Fig. 1D**). In addition, we associated these proteins with pathways, AMS symptom phenotypes, and clinical indexes to dissect the function of these proteins in AMS with the aim of redefining AMS with proteins and clinical indexes (**Fig. 1E**). Our study illuminates potentially important pathogenesis-related proteins, robust therapeutic targets, and predictive and diagnostic biomarkers of AMS with the aim of promoting an improved understanding and the redefinition of AMS.

**Figure 1.**
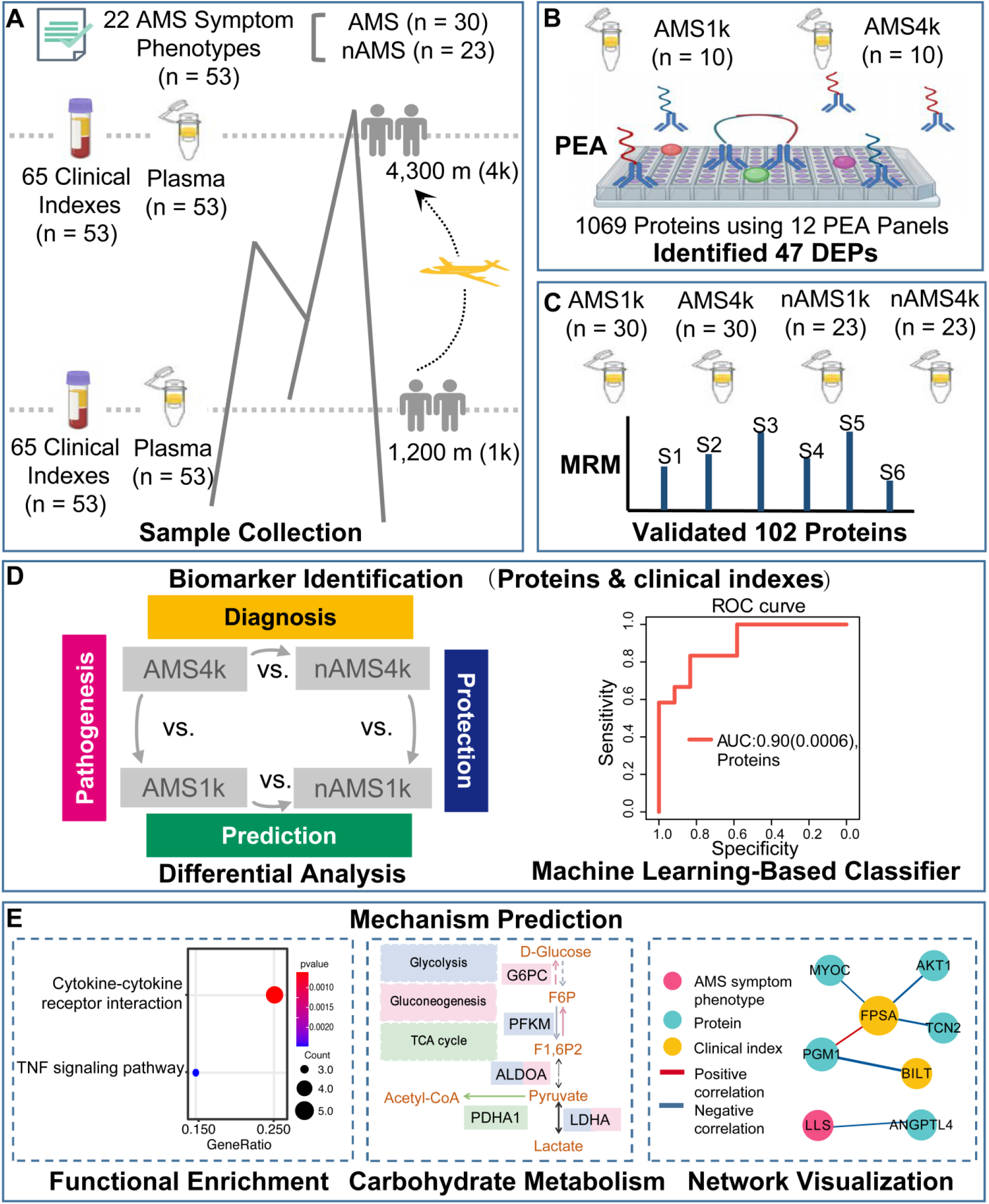
Study Design. (A) Sample collection. We recruited 53 subjects, collected plasma samples and recorded clinical indexes at plain (1 km) and high altitudes (4 km), and recorded 22 AMS symptom phenotypes at high altitude. According to the Louise Lake Score, the participants were separated into 30 individuals with AMS (AMS) and 23 individuals without AMS (nAMS). (B) At the discovery stage, PEA technology identified 47 plasma differentially expressed proteins (DEPs) between the 10 AMS4k and 10 AMS1k groups. (C) At the validation stage, MRM technology validated 102 proteins. (D) Differential analysis and machine learning-based classifiers with proteins and/or clinical indexes were used in the pathogenesis (pink background, comparison between the AMS4k and AMS1k groups), protection (dark blue background, comparison between the nAMS4k and AMS1k groups), prediction (dark green background, comparison between the AMS1k and nAMS1k groups), and diagnosis (yellow background, comparison between the AMS4k and nAMS4k groups) comparisons. (E) Functional enrichment analysis and network analysis were performed to illustrate the mechanism.

## Results

### Study design for exploring AMS

We recruited a cohort of 53 individuals in this study and collected 106 plasma samples (53 pairs) from the participants on the plain and at high altitude for 1-4 days. All the participants completed a questionnaire, which included 22 AMS symptom phenotypes (**Supplementary Table S1**), when they were at high altitude prior to blood collection. Among the tested symptom phenotypes, headache and 5 other symptom phenotypes were used to calculate the Louise Lake Acute Mountain Sickness Score (see Methods). To perform comprehensive proteomic profiling of AMS, we identified and quantified 1069 proteins in 20 plasma samples using PEA technology at the discovery stage and 102 proteins in 106 plasma samples using MRM technology at the validation stage. The discovery stage included two groups: individuals with AMS at high altitude (AMS4k) and the same individuals on the plain (AMS1k). The validation stage included four groups: individuals with AMS at high altitude (AMS4k), the same individuals on the plain (AMS1k), individuals without AMS at high altitude (nAMS4k), and the same individuals on the plain (nAMS1k).

### PEA-based identification of proteins involved in AMS at the discovery stage

At the discovery stage, we measured 1069 proteins in 20 plasma samples (10 AMS1k and 10 AMS4k) of 10 male individuals with AMS to explore the pathogenesis and therapeutic targets of AMS using 12 PEA panels (**Supplementary Table S2**). The average age and LLS score at high altitude were 18.8 and 5.2, respectively. Strict quality control identified 887 proteins (see Methods). Based on a q-value less than 0.05, we identified 47 differentially expressed proteins (DEPs), which included 40 upregulated and 7 downregulated proteins (**Fig. 2A** and **Supplementary Table S3**). ANGPTL4, MMP3, and FGF23 were upregulated in AMS4k, whereas CA1, CA2, and the chemokine CCL2 were downregulated (**Fig. 2A**).

**Figure 2:**
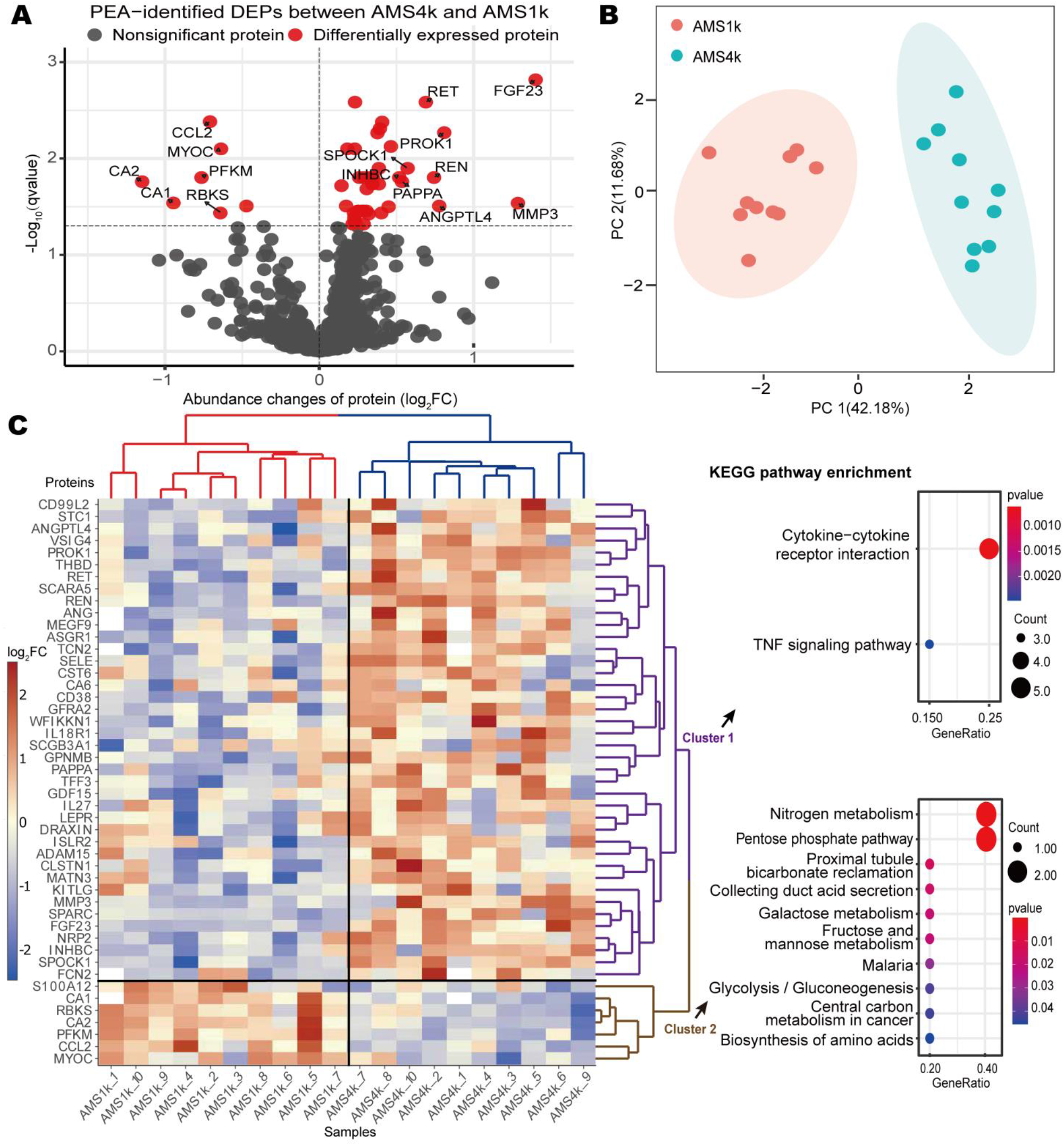
PEA-identified proteins and biological functions involved in AMS. (A) Volcano plot of PEA-identified DEPs between the AMS4k and AMS1k groups. Proteins with no statistical significance (q-value ≥ 0.05, gray dots) and DEPs (q-value < 0.05, red dots) with absolute value of log2 FC larger than 0.5 are labeled with gene symbols. (B) PCA plot based on PEA-identified DEPs. The AMS1k (light red dots) and AMS4k (sky-blue dots) samples were well separated by PCA using the PEA-identified DEPs without missing values (n = 42). (C) Abundance heatmap and KEGG pathway enrichment diagram of the PEA-identified DEPs. The scaled abundance heatmap of DEPs between the AMS4k and AMS1k groups per sample is shown on the left side. The hierarchical cluster analysis in the heatmap showed that the 47 DEPs could clearly distinguish the AMS4k group (red hierarchical tree) and AMS1k group (blue hierarchical tree), and the DEPs could be separated into cluster 1 (purple hierarchical tree) and cluster 2 (brown hierarchical tree). Dot plots of the KEGG enrichment of the two clusters with p-values less than 0.05 are presented on the right side. Cytokine-cytokine receptor interactions and the TNF signaling pathway were enriched in cluster 1 (purple fonts), and nitrogen metabolism and the pentose phosphate pathway were mainly enriched in cluster 2 (brown fonts).

The DEPs were sufficiently powered to clearly distinguish the AMS1k and AMS4k groups. A principal component analysis (PCA) showed that these proteins clearly distinguished the AMS4k group from the AMS1k group, and the first principal component accounted for 42.18% of the variance (**Fig. 2B**). A clustering analysis also showed clear separation between the two groups (**Fig. 2C**). Accordingly, we considered the 47 DEPs a primary clue for exploring the therapeutic targets of and protective, predictive, and diagnostic biomarkers for AMS.

### Identification of biological functions involved in AMS at the discovery stage

By performing a functional enrichment analysis of the upregulated and downregulated proteins, we identified pathways and Gene Ontology (GO) terms related to AMS. Energy metabolism pathways, such as nitrogen metabolism, the pentose phosphate pathway, and glycolysis/gluconeogenesis, were significantly downregulated in cluster 2, whereas cytokine-cytokine receptor interaction and TNF signaling pathways were significantly upregulated in cluster 1 (**Fig. 2C, Supplementary Table S4**). The GO enrichment analysis revealed that cell growth and response to stimuli, such as response to hypoxia and cAMP, were upregulated in cluster 1 (**Supplementary Fig. S1**), whereas bicarbonate transport, monocyte chemotaxis, and monosaccharide catabolic processes were downregulated in cluster 2 (**Supplementary Fig. S1**).

Carbonic anhydrase (CA) family proteins, including CA1, CA2, and CA6, are actively involved in AMS via the carbonate dehydratase process of the nitrogen metabolism pathway. In total, three types of cytokines, namely, the IL6 receptor family (LEPR), IL-1 receptor family (IL18R1), and TGF-β family (GDF15 and INHBC), which are involved in the cytokine-cytokine receptor interaction pathway, were upregulated. The results also revealed the upregulation of IL18R1, SELE, and MMP3, which are involved in the TNF signaling pathway. These findings indicate that immunological and inflammatory responses and energy metabolism are actively involved in the pathogenesis of AMS.

### Validation via Multiple Reaction Monitoring

To confirm the association between PEA-identified DEPs and AMS, we validated 102 proteins in 106 samples and 4 groups (AMS1k, AMS4k, nAMS1k, and nAMS4k) using an MRM MS-based proteome platform. No significant differences in age, height, or weight were found between the individuals with and without AMS, whereas the LLS scores at high altitude were significantly lower in the individuals without AMS than in the individuals with AMS (0.696 ± 0.703 vs. 4.3 ± 1.49, p-value < 0.001) (**Table 1**). We extended the 47 proteins to 102 proteins by adding 55 proteins involved in several pathways, such as the pentose phosphate and glycolysis pathways. Finally, we profiled 102 proteins (538 fragment ions) in 53 paired plasma samples (30 AMS4k, 30 AMS1k, 23 nAMS4k, and 23 nAMS1k) from 53 male Chinese Han subjects using MRM technology. The comparisons between the AMS4k and AMS1k, nAMS4k and nAMS1k, AMS1k and nAMS1k, and AMS4k and nAMS4k groups could be used to illustrate the pathogenesis, protection, prediction, and diagnosis of AMS, respectively (**Fig. 1D**).

**Table 1.** Baseline characteristics of the 53 subjects.

|  | nAMS<br>(n = 23) | AMS<br>(n = 30) | p-value |
| --- | --- | --- | --- |
| **Sex** |  |  |  |
| Male | 23 (100%) | 30 (100%) | NA |
| Female | 0 (0%) | 0 (0%) |  |
| **Age (years)** |  |  |  |
| Mean (SD) | 19.0 (1.30) | 19.3 (1.23) | 0.528 |
| Median [Min, Max] | 19.0 [17.0, 22.0] | 19.0 [17.0, 22.0] |  |
| **Height (cm)** |  |  |  |
| Mean (SD) | 171 (4.70) | 173 (5.47) | 0.157 |
| Median [Min, Max] | 170 [162, 180] | 173 [165, 185] |  |
| **Weight (kg)** |  |  |  |
| Mean (SD) | 63.1 (5.05) | 65.9 (7.00) | 0.0996 |
| Median [Min, Max] | 62.0 [54.0, 72.0] | 66.0 [53.0, 84.0] |  |
| **LLS** |  |  |  |
| Mean (SD) | 0.696 (0.703) | 4.3 (1.49) | < 0.001 |
| Median [Min, Max] | 1 [0, 2] | 4 [3, 9] |  |

#### Key proteins involved in the pathogenesis of AMS

The comparison between the AMS4k and AMS1k groups could be used to identify AMS pathogenesis-related proteins. We confirmed that 23 out of 47 proteins (49%) exhibited changing trends that were consistent with those found for the PEA-identified DEPs (**Supplementary Table S3**). Among these proteins, the MATN3, MYOC, RET, and S100A12 proteins were significantly different (**Fig. 3A**), which validates their potential significant involvement in the response to AMS. Myocilin, which is encoded by MYOC, is involved in cytoskeletal function. Matrilin 3 (MATN3) promotes the expression of HIF-1α(Q. Liu et al. 2018). S100A12 is a calcium-, zinc- and copper-binding protein that plays a prominent role in the regulation of inflammatory processes and the immune response(Pietzsch and Hoppmann 2009). RET, a transmembrane receptor and member of the tyrosine-protein kinase family of proteins, plays a role in cell differentiation, growth, migration, and survival. Notably, RET showed an opposite changing trend between nAMS4k and nAMS1k, although this difference was not statistically significant (**Fig. 3A**). In addition, the HIF-1 signaling pathway, glycolysis/gluconeogenesis, and carbon metabolism were enriched in DEPs identified by MRM (**Supplementary Fig. S2A and Supplementary Table S4**).

**Figure 3:**
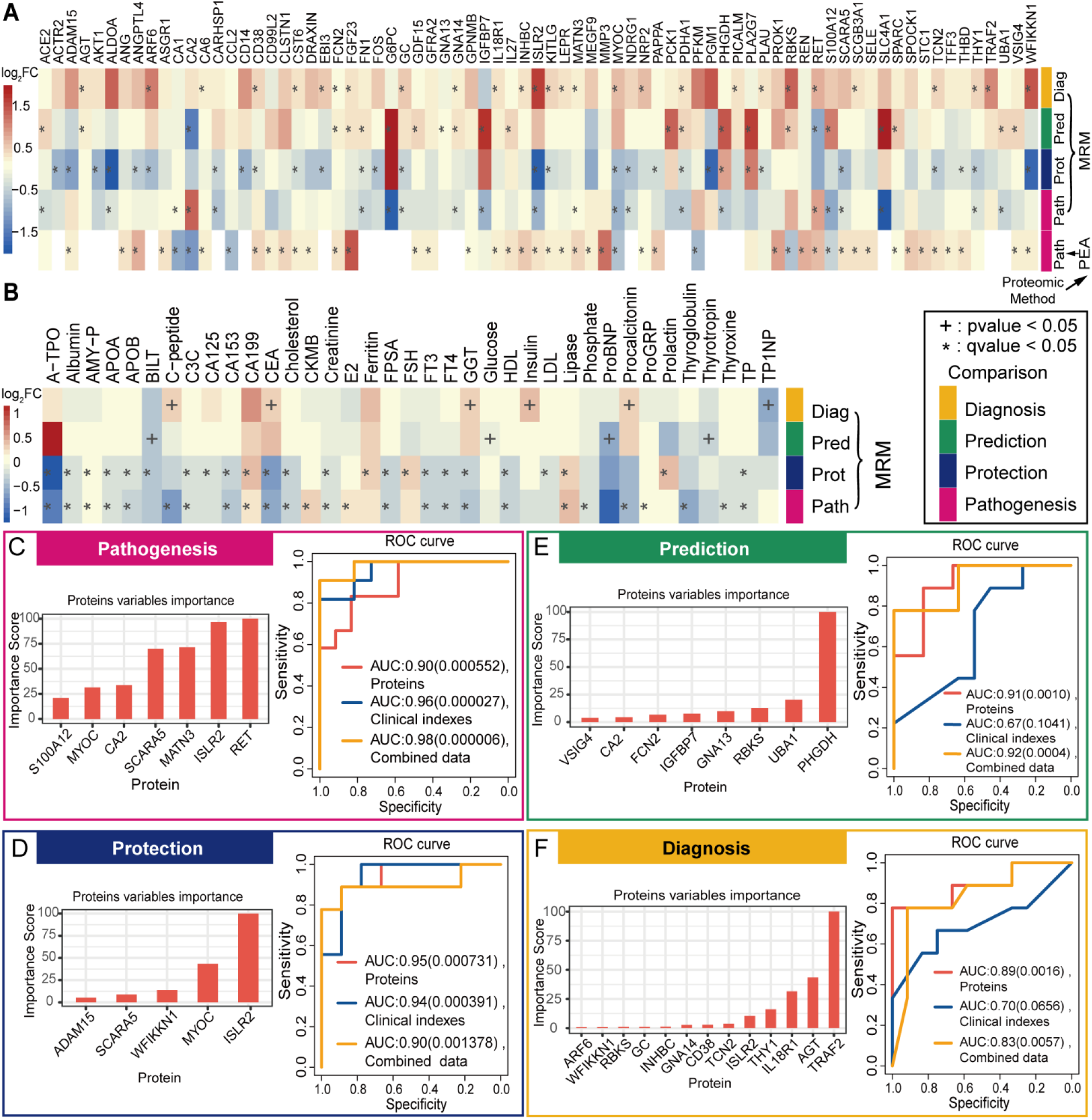
Differential analysis and classification with MRM-identified DEPs and clinical indexes. Differential analysis and machine learning were performed with the identified proteins and clinical indexes in 4 comparison groups (pathogenesis: AMS4k and AMS1k, protection: nAMS4k and nAMS1k, prediction: AMS1k and nAMS1k, and diagnosis: AMS4k and nAMS4k). (A) Heatmap of the log2FC of 75 PEA- or MRM-identified DEPs in the 5 comparison groups based on PEA or MRM. (B) Heatmap of the log2FC of 37 differential clinical indexes involved in the 4 comparison groups. The heatmap depicts the regulation trends of each protein or clinical index with the legend at the left of the figure. The proteins and clinical indexes were labeled with * (q-value < 0.05) and + (p-value < 0.05). (C) ROCs of the classification models with proteins (orange curve), clinical indexes (blue curve), or combined data (yellow curve) and bar plot of important proteins (orange bar) between AMS4k and AMS1k (pathogenesis, pink box). All the pathogenesis models with three types of features exhibit high accuracy with statistical significance, as demonstrated by the AUCs and p-values. (D) ROCs of the classification models with proteins (orange curve), clinical indexes (blue curve), or combined data (yellow curve) and bar plot of important proteins (orange bar) between the nAMS4k and nAMS1k groups (protection, dark blue box). All protection models with three types of features show high accuracy with statistical significance. (E) ROCs of the classification models with proteins (orange curve), clinical indexes (blue curve), or combined data (yellow curve) and bar plot of important proteins (orange bar) between the AMS1k and nAMS1k groups (prediction, dark green box). The prediction models with proteins and combined data show high accuracy. (F) ROCs of the classification models with proteins (orange curve), clinical indexes (blue curve), or combined data (yellow curve) and bar plot of important proteins (orange bar) between the AMS4k and nAMS4k groups (diagnosis, yellow box). The diagnosis models with proteins and combined data exhibit high accuracy with statistical significance.

#### Protective proteins for AMS

The comparison between the nAMS4k and nAMS1k groups could be used to explore protective proteins and could shed light on the pathophysiological mechanism and prevention of AMS. We identified 29 protective DEPs between the nAMS4k and nAMS1k groups **(Supplementary Table S3**). Overall, the trends obtained for ADAM15, CD38, CST6, KITLG, and THBD between these two groups were the opposite of those obtained for these proteins between the AMS4k and AMS1k groups, as revealed by both PEA and MRM (**Fig. 3A**), which indicates their potential protective role in preventing AMS and rapid acclimation to high altitude. Among these proteins, ADAM15 exhibited the highest degree of downregulation. In addition, the downregulated proteins CD38 and KITLG are involved in the hematopoietic cell lineage (**Supplementary Fig. S2B and Supplementary Table S4**), and KITLG is induced by HIF-1α under hypoxia in cancer cells(Han et al. 2008) and plays a role in hematopoiesis and cell migration.

#### Predictive biomarkers of AMS

Predicting the occurrence of AMS at high altitudes is difficult but has great value for individuals on plains. The comparison between the AMS1k and nAMS1k groups could identify predictive biomarkers. We identified 23 DEPs between the AMS1k and nAMS1k groups (**Supplementary Table S3**), and 17 of these DEPs showed the same trend as that obtained between the AMS4k and nAMS4k groups, showing the robustness of these proteins (**Fig. 3A**). Filtering based on absolute value of log2 FC larger than 0.5 resulted in 6 upregulated proteins, PCK1, PHGDH, RBKS, S100A12, SLC4A1, and SPARC. Among these DEPs, PCK1 and RBKS are involved in the metabolism of carbohydrates. These results indicate that these proteins are candidate predictive biomarkers of AMS.

#### Diagnostic biomarkers of AMS

The comparison between the AMS4k and nAMS4k groups could be used to discover diagnostic biomarkers of AMS. We identified 40 DEPs between the AMS4k and nAMS4k groups, and 28 of these proteins did not significantly differ between the AMS1k and nAMS1k groups (**Supplementary Table S3**). After filtering based on an absolute value of log2 FC larger than 0.5, 10 upregulated proteins (ARF6, EBI3, GC, ISLR2, MYOC, NRP2, RBKS, RET, TRAF2, and WFIKKN1) remained (**Fig. 3A, Supplementary Table S3**).

Among these proteins, EBI3, RET, and WFIKKN1 showed opposite regulation trends compared with those obtained from the comparison between the AMS1k and nAMS1k groups (**Fig. 3A**), which shows their higher possibility of being diagnostic biomarkers. Notably, RET exhibited the same significant regulation trends between the AMS4k and AMS1k groups, indicating its active role in the pathogenesis of AMS. ISLR2 is involved in axon guidance in brain development, and ISLR2 deficiency leads to severe hydrocephalus in mice(Abudureyimu et al. 2018). ISLR2 also shows a genetic interaction with RET(Mandai et al. 2009).

### High AMS classification accuracy

Four tree-based XGBoost machine learning models showed good performance in distinguishing AMS4k from AMS1k, nAMS4k from nAMS1k, AMS4k from nAMS4k, and AMS1k from nAMS1k. We used 10-fold cross-validation and the DEPs measured by MRM assays to enhance the robustness and performances of the models. The area under the curve (AUC) of the pathogenesis model with a panel of 7 proteins (RET, ISLR2, MATN3, SCARA5, CA2, MYOC, and S100A12) to distinguish AMS1k and AMS4k was 0.90 (p-value < 0.001, **Fig. 3C**). Notably, RET, MYOC, MATN3, and S100A12 were also candidate pathogenesis-related proteins via differential abundance analysis (**Fig. 3A**). MYOC, a type of secreted glycoprotein that participates in cell adhesion, cell-matrix adhesion, cytoskeleton organization, and cell migration, was reported to be reduced after incubation under hypoxia in trabecular meshwork cells and astrocytes(Obazawa et al. 2004).

The AUC of the protection model with 5 proteins (ISLR2, MYOC, WFIKKN1, SCARA5, and ADAM15) that distinguished nAMS1k and nAMS4k was 0.95 (p-value < 0.001, **Fig. 3D**), which indicated that these proteins may be involved in attenuating the symptoms of AMS in individuals without AMS at high altitudes. The responses of individuals with and without AMS to high altitude at the protein level were notably different. Notably, ADAM15 was identified as an essential candidate protective protein in the differential abundance analysis (**Fig. 3A**) and could therefore be a target for preventing and treating AMS.

The AUC of the prediction model distinguishing AMS1k and nAMS1k with 8 proteins (PHGDH, UBA1, RBKS, GNA13, IGFBP7, FCN2, CA2, and VSIG4) was 0.91 (p-value = 0.001, **Fig. 3E**). PHGDH and RBKS were also identified as candidate predictive biomarkers via differential abundance analysis, which suggested that these proteins likely serve as predictive biomarkers to evaluate the occurrence of AMS in individuals before their ascension to high altitude. In particular, PHGDH, which is involved in glucose/energy metabolism, had a 4-fold higher weight than other proteins in the model.

The AUC of the diagnosis model distinguishing AMS4k and nAMS4k with 13 proteins (TRAF2, AGT, IL18R1, THY1, ISLR2, TCN2, CD38, GNA14, INHBC, GC, RBKS, WFIKKN1, and ARF6) was 0.89 (p-value < 0.01, **Fig. 3F**), which indicated the potential translational value of these proteins as diagnostic biomarkers of AMS. TRAF2, AGT, and IL18R1 were the top 3 weighted proteins in this model. TRAF2, GC, and WFIKKN1 were also identified as candidate diagnostic biomarkers via differential analysis. TRAF2, a TNF receptor-associated factor, is associated with signal transduction from members of the TNF receptor superfamily and apoptosis. AGT, which is expressed in the liver, is involved in the maintenance of blood pressure, body fluid, and electrolyte homeostasis. IL18R1 is the receptor of the proinflammatory cytokine IL18. GC, a transporter of plasma metabolites binding to vitamin D, is reduced under hypoxia(Ma et al. 2012), which is consistent with our result that GC was downregulated in both individuals with AMS and individuals without AMS after ascending to high altitude.

Accordingly, we built robust models that distinguished these groups well and provided a panel of candidate biomarkers with promising translational value for each scenario.

### Key clinical indexes of AMS

The identification of key clinical indexes perturbed by AMS could shed light on our understanding and redefinition of AMS. Overall, 18 clinical indexes, such as A-TPO, albumin, C3C, FT3, and lipase (**Supplementary Fig. S3A and Fig. S3B**), showed significant differences between the AMS4k and AMS1k groups and between the nAMS4k and nAMS1k groups (**Fig. 3B, Supplementary Table 5**). Among these indexes, lipase, AMY-P, and CA199 were upregulated (**Supplementary Fig. S3A**). In addition, C-peptide, CKMB, E2, phosphate, proGRP, procalcitonin, thyroglobulin, and thyroxine showed significant differences between the AMS4k and AMS1k groups but not between the nAMS4k and nAMS1k groups, which reveals that these clinical indexes are affected by AMS (**Fig. 3B**). BILT, CA125, ferritin, FSH, LDL, and prolactin showed significant differences between the nAMS4k and nAMS1k groups but not between the nAMS4k and nAMS1k groups. Notably, BILT was upregulated in individuals who stayed at a high altitude for one month(Li et al. 2021). C3C, FT3, and TP1NP show significant differences between the Han and Tibetan populations(Jia et al. 2020).

BILT, glucose, proBNP, and thyrotropin were found to show significant differences between the AMS1k and nAMS1k groups (**Supplementary Fig. S3C**). TP1NP was significantly downregulated in the AMS4k group compared with the nAMS4k group, whereas C-peptide, CEA, GGT, insulin, and procalcitonin were significantly upregulated (**Supplementary Fig. S3D**). However, C-peptide, CEA, GGT, and procalcitonin were also significantly downregulated in the AMS4k group compared with the AMS1k group (**Fig. 3B**). The opposite trends between the two comparisons indicated the potentially remarkable roles of C-peptide, CEA, GGT, and procalcitonin in the pathogenesis of AMS. Procalcitonin is an inflammatory biomarker that is found at extremely low levels in the periphery of healthy individuals but increased by inflammatory mediators(Vijayan et al. 2017), which suggests the probable role of inflammation in the occurrence of AMS. Collectively, the association between AMS and multiple clinical indexes indicates that AMS is a complex and systemic disease.

We built robust models to distinguish AMS4k from AMS1k, nAMS4k from nAMS1k, AMS4k from nAMS4k, and AMS1k from nAMS1k using the clinical indexes identified from each differential analysis by 10-fold cross-validation. We found very high classification accuracy (AUC = 0.96, p-value < 0.0001) in the pathogenesis model distinguishing AMS1k and AMS4k with 19 clinical indexes (such as A-TPO, C-peptide, phosphate, E2, CKMB, and thyroxine, **Fig. 3C, Supplementary Fig. S4A**). The AUC of the protection model distinguishing nAMS1k and nAMS4k with a panel of 16 clinical indexes (such as FT3, ferritin, FPSA, creatinine, C3C, and FSH) was 0.94 (p-value < 0.001, **Fig. 3D, Supplementary Fig. S4A**), and the AUCs of the prediction and diagnosis models distinguishing AMS1k and nAMS1k with 3 clinical indexes (proBNP, BILT, and thyrotropin) and ASM4k and nAMS4k with 4 clinical indexes (procalcitonin, TP1NP, C-peptide, and insulin) were 0.67 and 0.7, respectively (p-values > 0.05, **Fig. 3E and Fig. 3F**). It appears that clinical indexes showed weaker performance than proteins for the prediction and diagnosis of AMS. A-TPO, which exhibited the highest weight in the AMS4k group compared with the AMS1k group (**Supplementary Fig. S4A**), is reportedly higher in hyperemesis gravidarum than in nonpregnant controls(Panesar et al. 2006), which indicates its association with vomiting.

Models were also established using a combination of clinical indexes and protein data to explore candidate biomarkers for AMS. The AUCs of the pathogenesis and protection models obtained using proteins and clinical indexes were 0.98 (p-value < 0.00001, **Fig. 3C**) and 0.9 (p-value < 0.01, **Fig. 3D**) between AMS4k and AMS1k and nAMS4k and nAMS1k, respectively. A-TPO continued to exhibit the top weight between the AMS4k and AMS1k groups, whereas RET, MYOC, and MATN3 were also selected from the combined data **(Supplementary Fig. S4B**). The classification performances of the pathogenesis (**Fig. 3C**) and protection (**Fig. 3D**) models obtained with proteins alone were comparable to those of the models obtained with the combined data.

In the prediction and diagnosis models, the classification performance obtained with protein alone and the combined data was similar. Notably, less features remained in the prediction model obtained with proteins alone than in those obtained with the combined data (**Fig. 3E, Fig. 3F, Supplementary Fig. S4B**). In addition, all important features remained in the diagnosis model obtained with the combined data were proteins. Accordingly, we hypothesized that proteins are better choices for the prediction and diagnosis of AMS than clinical indexes.

#### Changes in carbohydrate metabolism between individuals with and without AMS

Based on the dysregulation of PFKM and RBKS identified in the PEA assay (**Fig. 3A**), we extended the protein panel used in the MRM assay to comprehensively profile energy metabolism, particularly gluconeogenesis, glycolysis, and the tricarboxylic acid cycle (TCA cycle) (**Fig. 4, Supplementary Fig. S5B**) Individuals with AMS showed higher utilization of gluconeogenesis than individuals without AMS on the plain. The two key enzymes of gluconeogenesis, G6PC and PCK1, presented higher abundance in the AMS1k group than in the nAMS1k group (**Fig. 4 and Supplementary Fig. S5B**). Notably, G6PC is a subset of glucose-6-phosphatase catalyzing the hydrolysis of D-glucose 6-phosphate (G6P) to D-glucose, and PCK1 is a rate-limiting enzyme of gluconeogenesis.

**Figure 4:**
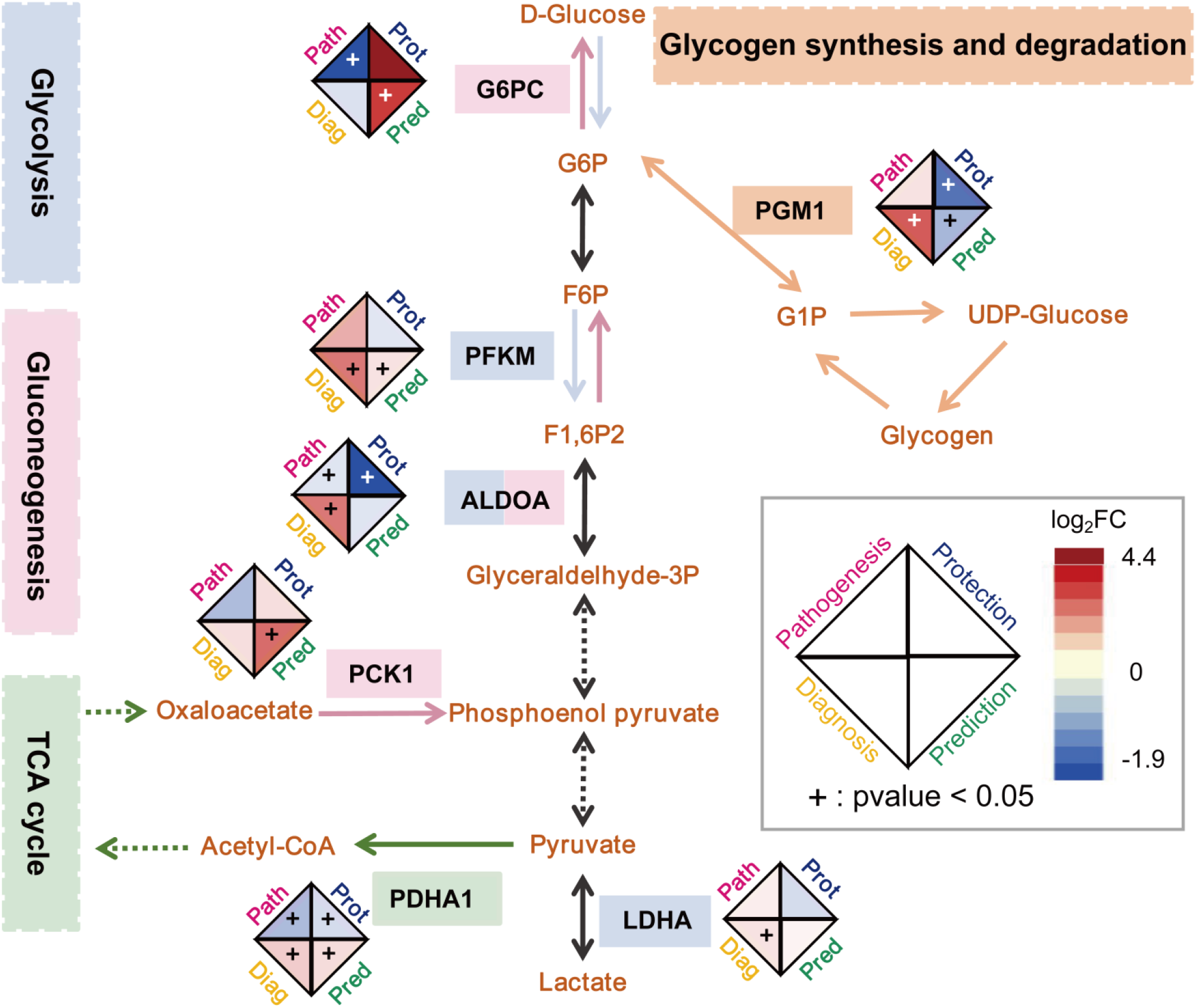
Changes in proteins involved in carbohydrate metabolism in AMS. The log2FC of proteins in glycolysis (light purple background and arrows), gluconeogenesis (light pink background and arrows), the TCA cycle (light green background and arrows), and glycogen synthesis and degradation (light orange background and arrows) in the following 4 comparison groups are shown in the diamond shape: pathogenesis (short for path, pink background), protection (prot, dark blue background), prediction (pred, dark green background), and diagnosis (diag, yellow background). Carbohydrate metabolism was dysregulated in individuals after ascension to high altitude.

Individuals with AMS may exhibit higher utilization of glycolysis than individuals without AMS at high altitude, and this hypothesis is supported by the following two points. First, the higher level of PFKM, a key rate-limiting enzyme in glycolysis, found in AMS4k than in nAMS4k would lead to the production of more fructose 1,6-bisphosphate and thereby the stimulation of glycolysis. Second, individuals with AMS showed higher levels of LDHA and ALDOA than individuals without AMS at high altitude. LDHA and ALDOA are involved in both glycolysis and gluconeogenesis. However, the gluconeogenesis-related enzymes PCK1 and G6PC showed similar expression levels between AMS4k and nAMS4k (**Supplementary Fig. S5B**). Therefore, we hypothesized that glycolysis, rather than gluconeogenesis, was more active in the AMS4k group than in the nAMS4k group.

Individuals with AMS may have a more active TCA cycle than individuals without AMS on the plain and at high altitude, although inhibition of the TCA cycle was found in both groups of individuals after ascension to high altitude (**Fig. 4**). PDHA1, a subset of pyruvate dehydrogenase, was downregulated after ascension to high altitude in both individuals with AMS and those without AMS. The downregulation of PDHA1 inhibited the conversion between pyruvate and acetyl-CoA and thereby downregulated the TCA cycle (**Supplementary Fig. S5B**), but the level in individuals with AMS remained higher than that in individuals without AMS.

Individuals without AMS had a lower utilization of glycogen after ascension to high altitude for better acclimation, whereas individuals with AMS may consume more glycogen for glycolysis than individuals without AMS at high altitude. Both UDP-glucose and food-derived glucose can participate in glycogen synthesis(Adeva-Andany et al. 2016). Glycogen can degrade into glucose 1-phosphate (G1P). The reversible isomerization between G1P and G6P is catalyzed by PGM1. PGM1 deficiency leads to a failure to utilize glycogen as an energy source in both the liver and skeletal muscle(Preisler et al. 2013). In our study, PGM1 was downregulated in nAMS4k compared with nAMS1k, lower in AMS1k than in nAMS1k, and higher in AMS4k than in nAMS4k (**Supplementary Fig. S5B**).

In summary, individuals with AMS may exhibit stronger gluconeogenesis ability on the plain and higher utilization of glycogen and glycolysis at high altitude than individuals without AMS. Individuals without AMS may inhibit the utilization of glycogen for storage after ascension to high altitude. After ascension to high altitude, all individuals showed a decreased TCA cycle, whereas individuals with AMS showed a more active TCA cycle than individuals without AMS. We argue that different carbohydrate metabolism pathways play a key role in acclimation to high altitude.

### Association among proteins, AMS symptom phenotypes, and clinical indexes

Associating the 22 symptom phenotypes of AMS with the changes in key proteins and clinical indexes between the AMS4k and AMS1k groups (**Fig. 5, Supplementary Table S6**) could dissect the AMS symptom phenotypes at a molecular level and better redefine AMS using proteins and/or clinical indexes. A correlation analysis showed that LLS was negatively correlated with S100A12, APOB, and cholesterol (ρs < -0.44, p-value < 0.05) in the C1 cluster (**Fig. 5**). Furthermore, the downregulation of S100A12, SLC4A1, IGFBP7, APOB, A-TPO, phosphate, and cholesterol was correlated with multiple symptom phenotypes, such as fatigue, nausea, vomiting, and poor appetite (C2 in **Fig. 5**), which indicates their potential important roles in the development of AMS. Hypoxia together with elevated inorganic phosphate could enhance vascular smooth muscle cell osteogenic transdifferentiation(Mokas et al. 2016). SLC4A1 is associated with CO2 gas transport in erythrocytes(Toye et al. 2008).

**Figure 5:**
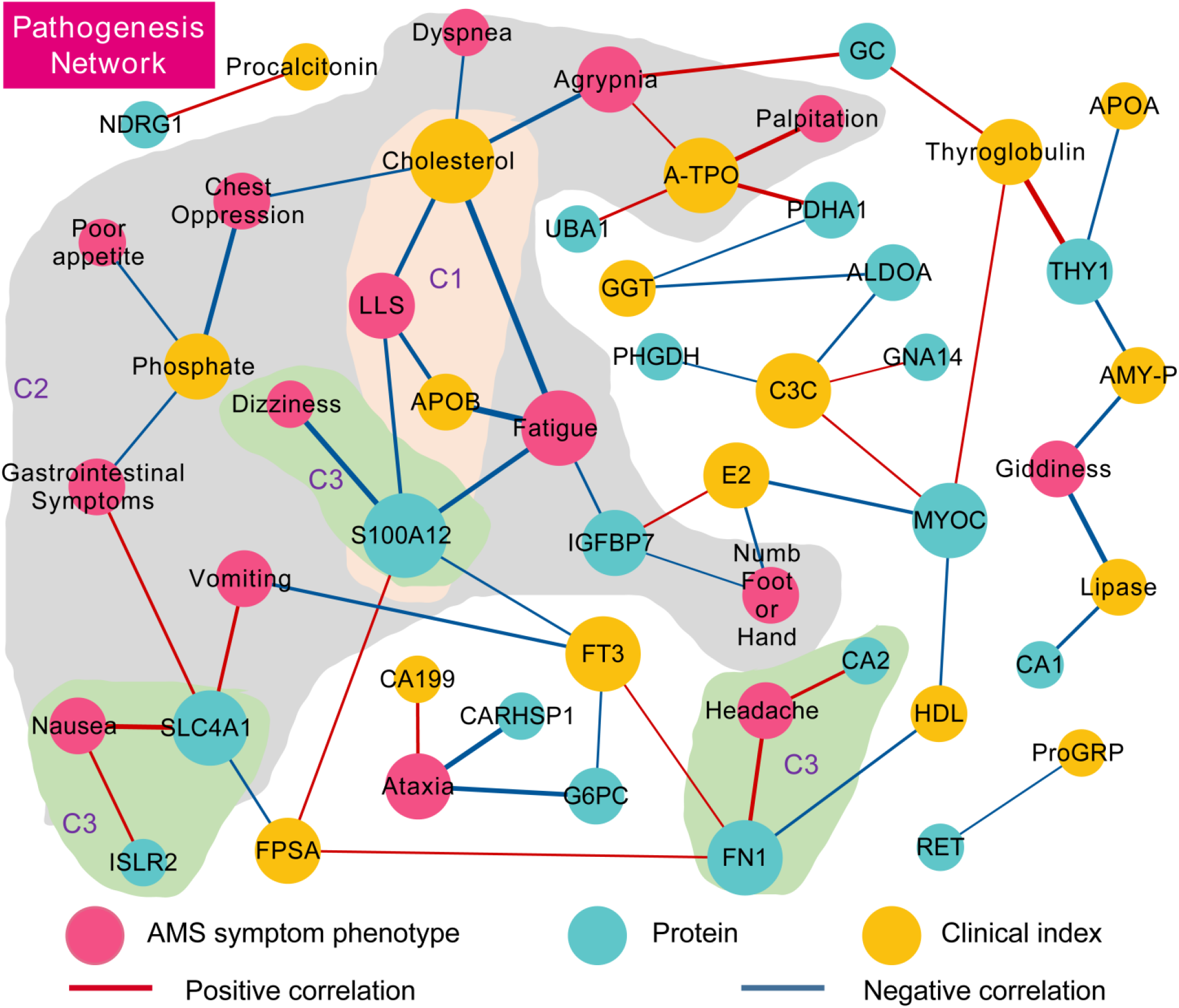
Network visualization of MRM-identified DEPs, clinical indexes and AMS symptom phenotypes in the pathogenesis comparison. The network of AMS symptom phenotypes (pink dots), DEPs (blue dots), and clinical indexes (yellow dots) was connected based on Spearman correlation coefficients and statistical significance. The edges showing a positive correlation (red line) and a negative correlation (blue line) with p-values less than 0.05 are shown. The thickness of the edges corresponds to the absolute value of the correlation coefficients. The size of the node is proportional to the number of edges from the node. Cluster C1 (light orange background area), cluster C2 (light green), and cluster C3 (light gray) are labeled C1, C2, and C3 (purple fonts), respectively. S100A12 was found to be associated with the LLS of AMS, and several proteins, such as CA2 and SLC4A1, which suggests their potential of being developed as novel clinical indexes.

Several AMS symptom phenotypes were correlated with proteins but not any clinical index (C3 in **Fig. 5**). For example, headache was positively correlated with CA2 and FN1 (ρs > 0.42, p-value < 0.05). Dizziness was negatively correlated with S100A12 (ρ = -0.51, p-value < 0.05), and nausea was positively correlated with ISLR2 and SLC4A1 (ρs > 0.41, p-value < 0.05). These results indicate opportunities for these proteins to be developed into novel clinical indexes to better redefine AMS.

The correlation analysis among clinical indexes, proteins, and AMS symptom phenotypes in 53 individuals on the plain or at high altitude revealed that several features could contribute to the prediction, pathogenesis, or diagnosis of AMS. No significantly different features between AMS1k and nAMS1k were found to be correlated with the AMS symptom phenotypes (**Supplementary Fig. S6A**). However, a connection between thyrotropin and the glucose/energy metabolism-related proteins PCK1, PHGDH, and IGFBP7 and the phospholipid catabolism-related protein PLA2G7 was found between AMS1k and nAMS1k. The SNPs of thyrotropin and PLA2G7 are associated with blood pressure variations and hypertension, respectively(Kokubo et al. 2006). Moreover, rather than clinical indexes, proteins, including FGF23, RET, IL18R1, and GNA14, were positively correlated with AMS symptom phenotypes, such as poor appetite, dyspnea, and lip cyanosis, between the AMS4k and nAMS4k groups (**Supplementary Fig. S6B**). In addition, C-peptide was positively correlated with 13 proteins, such as the candidate pathogenesis-related protein MYOC (ρ = 0.42, q-value < 0.05), the protective protein KITLG (ρ = 0.27, q-value < 0.05), and the diagnostic biomarker TRAF2 (ρ = 0.36, q-value < 0.05). Accordingly, C-peptide may be involved in the pathogenesis of AMS, whereas the clinical indexes investigated in our study did not perform better than proteins in the prediction and diagnosis of AMS.

## Discussion

We performed a systematic study of AMS using the plasma proteome and clinical indexes. Specifically, we profiled 1,069 proteins from 10 individuals with AMS on the plain and at high altitude using Olink’s PEA technology and identified 47 AMS-relevant proteins. PEA technology exhibits high sensitivity down to concentrations at the pg/mL range, which allows the detection of low-abundance proteins(Petrera et al. 2021). We validated 102 proteins using MS-based MRM technology in 53 individuals with or without AMS on the plain and at high altitude. The combination of PEA and MRM could largely avoid false positives, increasing the quality of the candidate biomarkers(Petrera et al. 2021). We then systematically analyzed the proteins and clinical indexes that may be involved in the pathogenesis, protection, prediction, and diagnosis of AMS to identify candidate therapeutic targets and biomarkers. We also discovered carbohydrate metabolism dysregulation in individuals with AMS and without AMS after ascension to high altitude. Moreover, we profiled 22 AMS symptom phenotypes and 65 clinical indexes in the same cohort and identified strong correlations among AMS-related proteins, phenotypes, and clinical indexes, which provides a basis for the precise redefinition of AMS using proteins and clinical indexes.

### RET may be highly involved in the development of AMS and could be a candidate therapeutic target and diagnostic biomarker for AMS

The PEA and MRM assays confirmed that plasma RET was significantly increased in individuals with AMS at high altitude compared with that found in these individuals on the plain (**Supplementary Fig. S5A**). However, RET was decreased in individuals without AMS at high altitude compared with the levels found in these individuals on the plain (**Supplementary Fig. S5A**). RET was identified as the most important variable in the machine learning-based model with high classifier accuracy (AUC = 0.9) for the AMS4k and AMS1k groups.

RET is correlated with a basic symptom of AMS. RET could regulate the survival and size of nociceptors that transmit information to the brain, leading to the sensation of pain(Golden et al. 2010; Kelman 2011). In our study, RET was positively correlated with headache (ρ = 0.35, p-value = 0.06, **Supplementary Table S6**). This result suggests that RET is likely involved in the pathophysiological mechanism of headache, a key phenotype of AMS.

RET could increase the promoter activity of CA9 and LDHA induced by HIF-1α under hypoxia(Takacova et al. 2014; Greer et al. 2012). CA9, which is homologous with CA1, CA2, and CA6, could maintain the homeostasis of the blood oxygen partial pressure and acid-base balance by regulating ventilation under hypoxia(Trupp et al. 1998; Tano et al. 2009). CA2, which was also selected by XGBoost, was identified as a candidate predictive biomarker for AMS in our study. Acetazolamide, an inhibitor of CA2 and CA9, is a class of drugs to prevent and treat AMS. These results suggest that RET likely also interacts with CA2. In addition, ISLR2, which exhibited the top weight in the machine learning model to distinguish nAMS4k and nAMS1k, is related to RET(Mandai et al. 2009). Remarkably, the selective inhibitors of RET selpercatinib and pralsetinib were recently approved for the treatment of RET-associated non-small cell lung cancer(Vivek Subbiah et al. 2018; V. Subbiah et al. 2018). In summary, RET could be a therapeutic target in the treatment of AMS.

In addition, proinflammatory programs are selectively activated by TRAF2 and TRAF6, which are associated with RET and PTC oncoproteins in papillary thyroid carcinoma(Wixted, Rothstein, and Eisenlohr 2012). TRAF2 was also selected by both differential analysis and XGBoost between AMS4k and nAMS4k as a candidate diagnostic biomarker. In addition, RET was positively correlated with lip cyanosis, an AMS symptom phenotype. Therefore, RET may be an essential diagnostic biomarker for AMS.

### TRAF2 is a candidate diagnostic biomarker for AMS

TRAF2 was selected via differential analysis and the XGBoost classifier with an AUC of 0.89 between AMS4k and nAMS4k (**Fig. 3F**). TRAF2 interacts with procaspase-12 and promotes the activation of caspase-12(Yoneda et al. 2001), which can transduce signals from IRE1s under ER stress conditions and lead to apoptosis(Nakagawa et al. 2000). Moreover, salidroside reportedly reduces TRAF2 to protect against hypoxia-induced liver injury by inhibiting ER stress-mediated apoptosis(Xiong et al. 2020). Accordingly, TRAF2 was identified as a candidate diagnostic biomarker for AMS, and salidroside was found to be a candidate drug for the treatment of AMS.

### The downregulation of S100A12 may not prevent AMS

The PEA and MRM assays confirmed that S100A12 was significantly decreased in individuals with AMS at high altitude compared with the levels found in these individuals on the plain (**Fig. 3A**). Among all the groups, the highest expression of S100A12 was found in the AMS1k group (**Supplementary Fig. S5A**). In addition, S100A12 was one of the key features in the machine learning-based pathogenesis model with high accuracy and was identified as a candidate predictive biomarker via differential analysis between the AMS1k and nASM1k groups. Moreover, S100A12 was negatively correlated with LLS, dizziness and fatigue. These findings indicate that S100A12 is highly involved in the pathogenesis of AMS.

The downregulation of S100A12 did not inhibit the inflammatory response induced by hypoxia. The inflammatory response was actively involved in AMS, as shown previously. The proinflammatory factor S100A12, a type of endogenous innate danger molecule, could provoke proinflammatory responses in endothelial cells(van Zoelen, Achouiti, and van der Poll 2011). The S100A12 levels are associated with increased levels of markers of pulmonary inflammation and hypoxia in patients undergoing cardiac surgery(Müller et al. 2014). In addition, the downregulation of S100A12 in aortic smooth muscle could reduce apoptosis(Das et al. 2012) but has no significant effect on inflammatory signaling in monocytes(Foell et al. 2013). Therefore, it appears that the downregulation of S100A12 likely reduces apoptosis but does not inhibit the inflammation induced by hypoxia in individuals with AMS after ascension to high altitude.

### The downregulation of ADAM15 may play a protective role in individuals without AMS

ADAM15 was identified as a candidate protective biomarker of individuals without AMS and was selected by the machine learning-based model with high accuracy to distinguish nAMS1k and nAMS4k. ADAM15 is involved in the response to hypoxia, proteolytic ectodomain processing of cytokines, cell adhesion signaling, and angiogenesis in endothelial cells(Horiuchi et al. 2003; Nishimi et al. 2019). In addition, the silencing of ADAM15 can inhibit the expression of proinflammatory cytokines in rheumatoid angiogenesis(Nishimi et al. 2019). Moreover, systemic proinflammatory cytokines are associated with the development of AMS and high-altitude pulmonary edema(B. Liu et al. 2017; Song et al. 2016). Proteins involved in the inflammatory response, such as FGF23, KITLG, and PLAU, are found at lower level in individuals without AMS than in those with AMS, but CCL2 was not significant in any of the compared groups (**Supplementary Fig. S5A**). This finding is consistent with the fact that CCL2 is independent of the AMS status(Colleen Glyde Julian et al. 2011). Importantly, adequate anti-inflammatory properties favor resistance to AMS(Colleen Glyde Julian et al. 2011). The candidate pathogenesis biomarker RET, which was higher in individuals with AMS than in those without AMS, was associated with proinflammation. In conclusion, subjects without AMS may rapidly acclimate to a high-altitude environment by downregulating ADAM15 and inflammation. ADAM15 could be a target in the treatment of AMS.

### PHGDH may be a candidate predictive biomarker for AMS

PHGDH was identified as a candidate predictive biomarker of AMS, which was the top-weighted protein identified by the XGBoost classifier with an AUC of 0.91 between AMS1k and nAMS1k. PHGDH is involved in glucose and energy metabolism. The inhibition of PHGDH could downregulate the NADPH levels, disorder mitochondrial redox homeostasis, and increase apoptosis under hypoxia(Samanta et al. 2016; Engel et al. 2020). The overexpression of PHGDH reduces hypoxia-induced cell death(Engel et al. 2020). Moreover, in our study, PHGDH was upregulated in individuals without AMS and downregulated in individuals with AMS to a similar level after ascension to high altitude (**Supplementary Fig. S5A**). Collectively, the results indicate that PHGDH may be a promising predictive biomarker for AMS.

### C-peptide may be associated with the pathogenesis and diagnosis of AMS

C-peptide was selected via both differential analysis and the classifier between the AMS4k and AMS1k and the AMS4k and nAMS4k groups with AUCs greater than 0.7 but not between the nAMS4k and nAMS1k groups. C-peptide, which is a polypeptide that connects two chains of proinsulin, was downregulated in AMS4k compared with AMS1k but was still higher in individuals with AMS than in those without AMS at high altitude. In addition, our study showed that C-peptide was positively correlated with proteins related to the pathogenesis, protection and diagnosis of AMS. Moreover, C-peptide could increase proliferation(Mughal et al. 2010) and activate anti-inflammation in endothelial cells(Luppi et al. 2008). C-peptide is reportedly elevated in 7 subjects with AMS after ascension to high altitude(Larsen et al. 1997), which is consistent with our result **(Fig. 3B**). However, an acclimatization study revealed that C-peptide was significantly decreased at 3,600 m compared with the sea level, but no significant difference was found between sea level and an altitude of 4,650 m and above, regardless of the AMS status(Hill et al. 2018). In our study, C-peptide showed the same downregulation trend in both individuals with and without AMS at high altitude compared with the levels found in these individuals on the plains. Taken together, the results show that C-peptide may be associated with the pathogenesis and diagnosis of AMS.

### Carbohydrate metabolism plays a key role in AMS

Individuals with AMS may have a more active TCA cycle than individuals without AMS in response to hypoxia and show enhanced glycolysis and increased utilization of glycogen compared with individuals without AMS at high altitude. In addition, individuals with AMS exhibit active gluconeogenesis on the plain. Regardless of whether the individuals suffered from AMS at high altitudes, individuals at high altitudes showed a reduced TCA cycle due to the hypoxic environment. Lu et al. reported that the TCA cycle and glycolysis are reduced in individuals without AMS but not in individuals with AMS after exposure to high altitude based on the downregulation of TCA-related enzymes (such as PDHA1) and glycolysis-related enzymes (such as ALDOA) in the AMS-resistant group(Lu et al. 2018). It appears that the balance between glycolysis and gluconeogenesis is relevant to AMS. These differences could aggravate the consumption of oxygen, leading to discomfort in individuals with AMS and comfort in individuals without AMS at high altitude. Additionally, an enzyme related to glycogenesis, PCK1, was identified as a candidate predictive biomarker for AMS in our study, and this finding highlights the potentially important role of gluconeogenesis in the prediction of AMS.

### Robust and applicable machine learning models for AMS

We built four robust machine-learning models to dissect the pathogenesis of AMS, screen therapeutic targets and identify protective, predictive and diagnostic biomarkers. Using only several proteins, these models maintained high accuracy (AUCs ≥ 0.9), which implies the applicable value of these models. In particular, we could screen individuals susceptible to AMS using predictive biomarkers to largely prevent the occurrence of AMS, which is undesirable to individuals who would like to ascend to high altitude. The pathogenesis and protection models obtained using clinical indexes exhibited high accuracy, whereas the prediction and diagnosis models established using clinical indexes did not perform as well as those obtained using proteins. Moreover, the prediction and diagnosis models obtained using both clinical indexes and proteins did not perform as well as those established using only proteins, and this finding indicates that these proteins are more suitable for the prediction and diagnosis of AMS than these clinical indexes, which shows their potency as novel clinical indexes.

### Precise redefinition of AMS based on proteins and clinical indexes

The redefinition of AMS based on proteins and clinical indexes will promote an improved understanding of AMS and precise treatment. Currently, the diagnosis of AMS mainly depends on the self-questionnaire LLS. Here, we propose a panel of candidate predictive biomarkers (such as PHGDH, UBA1, RBKS, GNA13, IGFBP7, CA2, and VSIG4) and a panel of candidate diagnostic biomarkers (such as TRAF2, AGT, IL18R1, ISLR2, GC, RBKS, and WFIKKN1) for AMS. In addition, C-peptide could be an assistant diagnostic biomarker for AMS identified via differential analysis, the machine learning model, and correlation analysis with multiple AMS-relevant proteins.

### Limitation and future work

In this study, we used 53 paired plasma samples from 53 individuals with robust statistical and machine learning-based models to comprehensively profile AMS utilizing PEA and MRM-based proteomic technology. To the best of our knowledge, although this is the largest cohort used in a study of AMS, we plan to further validate these biomarkers using another independent cohort and *in vivo* experiments to promote translational medicine.

### Conclusion

We systematically profiled the characteristics of AMS using two proteome technologies based on different principles, PEA and MRM, and 106 plasma samples. We validated that RET actively participates in the pathogenesis of AMS and has the potential to be a candidate therapeutic target. The downregulation of ADAM15 may play a role in preventing individuals from developing AMS. PHGDH is a candidate predictive biomarker, and TRAF2 is a promising diagnostic biomarker. Furthermore, we built robust machine learning-based diagnosis, prognosis, protection, and pathogenesis models with high classification accuracy and thereby validated the roles of these proteins in AMS. Individuals with AMS may exhibit more active gluconeogenesis on the plain than individuals without AMS and enhance the utilization of glycogen compared with individuals without AMS at high altitude. Additionally, we profiled the associations among 22 symptom phenotypes of AMS, 65 clinical indexes, and these proteins, and the findings indicate the possibility of redefining AMS based on proteins and clinical indexes instead of a self-questionnaire and shed light for obtaining an improved understanding of AMS, better acclimatization to a hypoxic environment, and the promotion of precision medicine for AMS.

## Materials and Methods

### Subjects

A total of 53 Han Chinese male subjects (aged 18-20 years) were recruited in this study. The exclusion criteria included having any health problems; having any known liver, lung, or cardiovascular disease; a history of migraine or head injury; smoking; and having been to altitudes > 2500 m or exposed to a hypobaric hypoxic environment within the last three months. Ethical approval was achieved from the Chinese PLA General Hospital ethical committee with the approval identifier S2019-035-01, and all protocols followed the established national and institutional ethical guidelines. All participants provided signed written informed consent.

### Evaluation of the AMS status at high altitude

The AMS status of 53 subjects was evaluated according to the LLS(Roach et al. 2018), a self-reported scoring standard, after ascension to high altitude (4,300 meters). Briefly, a four-point scale (asymptomatic = 0, mild = 1, moderate = 2, severe = 3) was used to quantify the degree of headache, gastrointestinal symptoms (poor appetite, nausea/vomiting), fatigue and dizziness (**Supplementary Table S1**). Subjects with severe headache but no other symptoms of AMS or an LLS greater than 2 were defined as individuals with AMS (AMS, n=30). Subjects with an LLS less than 3 and subjects without headaches were defined as individuals without AMS (nAMS, n=23). With the exception of the 6 symptom phenotypes used for calculating the LLS, 14 other AMS symptom phenotypes were also evaluated for further analysis (**Supplementary Table S1**).

### Experimental set-up

All subjects were transported to an altitude of 4,300 meters (4 km) from 1,200 meters (1 km) within 4 hours by plane. Peripheral venous blood samples were collected at both 1 km and 4 km. The AMS symptoms were evaluated after ascension to 4 km. Twenty paired plasma samples from 10 individuals with AMS at 1 km (AMS1k) and 4 km (AMS4k) were selected for identifying the protein expression profile using Olink’s PEA technology. Subsequently, 60 paired plasma samples from 30 individuals with AMS (including the subjects used for PEA) and 46 paired plasma samples from 22 individuals without AMS were used for validation.

### Sample collection and biochemical detection

Peripheral venous whole-blood samples of 53 subjects (30 AMS and 23 nAMS) were collected at an altitude of 1 km (AMS1k and nAMS1k) and after arrival at an altitude of 4 km (AMS4k and nAMS4k) for 1-4 days. Blood samples were collected in a semi-recumbent position from an anterior elbow vein by conventional venipuncture and placed in an EDTA-coated blood collection tube. Plasma was separated by centrifugation and stored in a 0.5-mL aliquot at -80°C until analysis. All samples from these subjects at both time points were also prepared for biochemical detection. Two milliliters of each plasma sample was used to assay 65 clinical indexes (**Supplementary Table S1**) using a hematology analyzer (cobs 6000; Roche, USA).

### Plasma proteome profiling and analysis

We analyzed 20 plasma samples from individuals with AMS using Olink’s PEA technology with 12 panels by iCarbonX (Shenzhen) Company Limited. These panels consisted of 6 disease panels (Cardiometabolic panel, Cardiovascular II panel, Cardiovascular III panel, Development panel, Neurology panel, Neuro Exploratory panel, Oncology II panel, and Oncology III panel) and 6 important biological progress panels (Immune Response panel, Cell Regulation panel, Immuno-Oncology panel, Inflammation panel, Metabolism panel, and Organ Damage panel), as indicated on the manufacturer’s website (Olink Proteomics, Uppsala, Sweden). Each PEA panel required 1 μL of plasma sample. The protein concentrations were finally normalized and transformed using internal and interplate controls to adjust for intra- and inter-run variation(Assarsson et al. 2014). More detailed information can be found in the panel-specific validation data documents (www.olink.com/downloads). The expression levels of proteins were represented as linear Normalized Protein eXpression (NPX), a relative quantification scale in arbitrary units. A complete list of all 1104 measured proteins (1069 unique proteins) can be found in the supplementary data (**Supplementary Table S2)**.

Proteins with coefficients of variation less than 0.3 and a missing data frequency less than 0.25 were used for further analysis. The values of undetected features were replaced with 1/10 of the minimum nonzero value. Paired t-tests or paired Welch’s t-tests were performed for statistical analyses based on homoscedasticity, and the p-values were corrected using Bonferroni-Hochberg (BH) corrections (q-value) for multiple comparisons. Proteins with q-values less than 0.05 were considered DEPs. PCA was performed using the DEPs without missing value via the package ggplot2(Gómez-Rubio 2017). The heatmap of DEPs was generated using the R package heatmaply(Galili et al. 2018) with scaled raw data.

### DEP validation by multiple reaction monitoring

MRM was applied to verify the 47 selected DEPs measured by PEA and 55 other pathway-related proteins at the validation stage, which consisted of 106 samples (30 individuals with AMS and 23 nAMS at 1 km and 4 km, respectively). A total of 538 transitions were selected to represent the 102 proteins. Unique peptides were selected from the peptides identified by TripleTOF 5600+ mass spectrometry (SCIEX) in the mixed plasma samples and the public library SRMAtlas. The MRM assay was performed using a QTRAP 6500+ mass spectrometer (SCIEX), and the quantitative method was constructed by the Analytics module in SCIEX OS software (version 2.0) (Beijing Qinglian Biotech Company Limited). By using BSA peptides for normalization, we obtained the standardized abundance value (intensity) and performed relative quantitation according to grouping. Transitions with missing value frequencies greater than 25% were removed. Further calculations and statistical analyses were conducted using paired t-test or paired Welch’s t-test for the self-control compared groups and t-test or Welch’s t-test for the other compared groups in R based on homoscedasticity. The p-values were corrected using BH (q-value) for multiple comparisons. Proteins with q-values less than 0.05 were considered DEPs. The heatmap of the intersection of MRM-validated DEPs selected via differential analysis in four compared groups was generated using the pheatmap package, and the violin plot was generated using ggplot2.

### Functional enrichment analysis

Gene ontology biological process (GOBP) and Kyoto Encyclopedia of Genes and Genomes (KEGG) enrichment analyses of the DPEs measured by PEA and MRM were performed using the Bioconductor R package clusterProfiler(Yu et al. 2012). The redundant GO terms were removed using the simplifying function (by = “p.adjust”, cutoff = 0.3). The statistical significance of the GO enrichment was tested using Benjamini and Hochberg with a cutoff q-value less than 0.05. All the KEGG analyses with p-values less than 0.05 were enriched and shown.

### Machine learning-based models

XGBoost, a boosted ensemble algorithm, was implemented, and 10-fold cross-validation was performed using MRM-based proteomic data and/or clinical indexes for the identification of biomarkers. The proteins used for prediction were selected at the validation stage based on a q-value less than 0.05. Clinical indexes were selected based on q-values less than 0.05 between the AMS4k and AMS1k groups and the nAMS4k and nAMS1k groups and by p-values less than 0.05 between the AMS1k and nAMS1k groups and the AMS4k and nAMS4k groups. The datasets between each comparison were divided into a training set (60%) and a test set (40%). XGBoost was performed using the caret package with the xgbTree method. Receiver operating characteristic (ROC) curves were generated to assess the AUC with the pROC package(Robin et al. 2011).

### Correlations among proteins, clinical indexes, and symptom phenotypes

Spearman correlation analyses among proteins, clinical indexes, and AMS symptom phenotypes were performed using package psych, and the results were visualized using Cytoscape software(Shannon et al. 2003). Samples with less than 75% of observations were eliminated during the correlation analysis. In addition, features observed in less than 25% of the samples were also deleted with the exception of the symptom phenotypes involved in LLS between the AMS4k and AMS1k groups and the AMS4k and nAMS4k groups. Additionally, features with the same value in each sample were also deleted. The p-values were corrected using BH (q-value) for multiple comparisons. Connections with p-values less than 0.05 were selected for network visualization between the AMS4k and AMS1k groups and the nAMS4k and nAMS1k groups because no connections showed q-values less than 0.05, whereas connections with q-values less than 0.05 were selected from the other two compared groups.

### Statistical analysis

For the baseline data, continuous variables are presented as the means ± standard deviations (SDs) and medians with interquartile ranges (IQRs), and the ordinal and nominal variables are presented as percentages. The NPX data generated from PEA were compared by paired t-test. The data generated by MRM were compared by paired and unpaired t-test or Welch’s t-test based on homoscedasticity. The clinical indexes were compared by t-test, Welch’s t-test, or Wilcoxon test (paired or unpaired) considering a normal distribution and homoscedasticity. The p-values were corrected using BH (q-value) for multiple comparisons. Proteins with q-values less than 0.05 were defined as DEPs. All statistical analyses were performed using R (version 4.0.2).

## Data Availability

The raw MRM proteomic data analyzed in this study are available at iProX with the corresponding dataset identifier PXD029063. The source code is freely available at https://github.com/Monica1227/AMS_biomarker.

## Availability of Data

The raw MRM proteomic data analyzed in this study are available at iProX(Ma J, et al. 2018) with the corresponding dataset identifier PXD029063. The source code is freely available at https://github.com/Monica1227/AMS_biomarker.

## Acknowledgments

We thank the participants who donated samples and all colleagues who contributed to this study during the sample collection.

## Author contributions

Conceptualization: K.H., Z.J.; Sample collection: J.S., X. Z. and Z.J.; Data analysis: J.Y., X.S.; Data interpretation: Z.J. and J.Y.; Writing: J.Y.; Editing: Z.J., J.Y. and K.H.. All authors read and approved the final manuscript.

## Ethics approval

Ethical approval was achieved from the Chinese PLA General Hospital ethical committee with the approval identifier S2019-035-01, and all protocols followed the established national and institutional ethical guidelines. All participants provided signed written informed consent.

## Supplementary files

**Figure S1:**
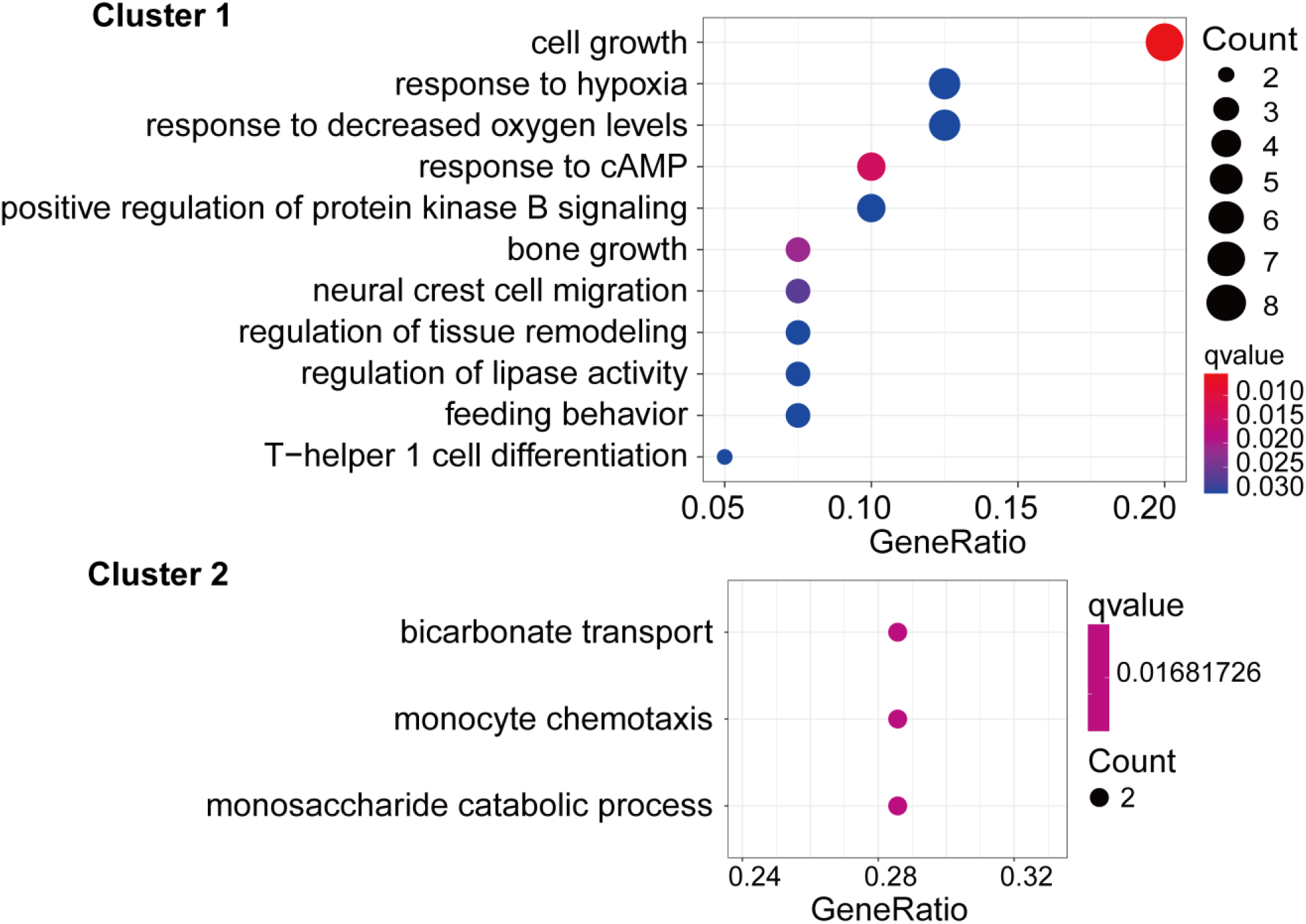
Dot plot of biological process-related gene ontology enrichment of the two clusters. Terms with a q-value less than 0.05 are shown. Cell growth and response to stimulus were mainly enriched in cluster 1 (up panel), and bicarbonate transport was mainly enriched in cluster 2 (down panel).

**Figure S2:**
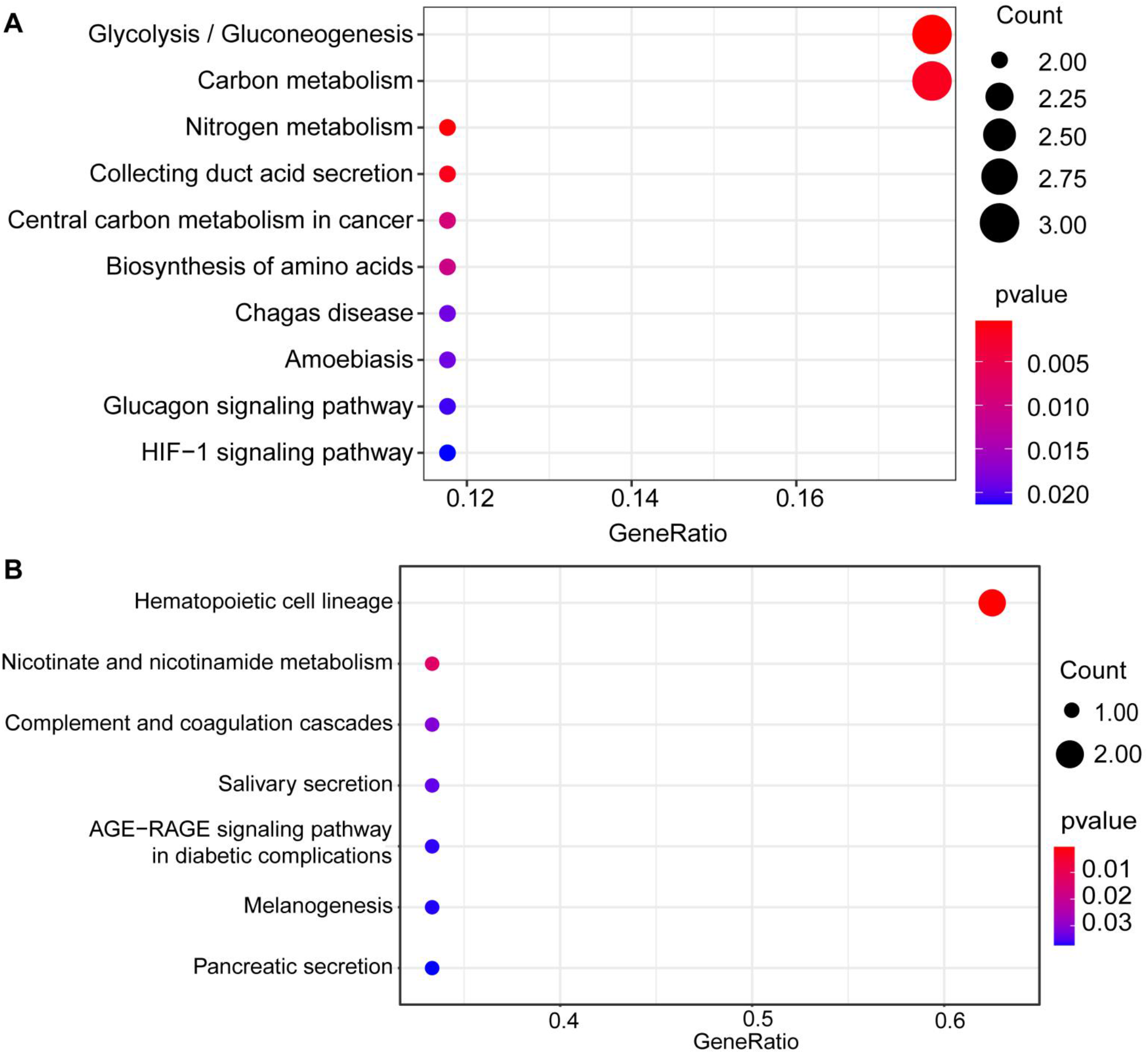
Dot plot of KEGG enrichment of MRM-identified DEPs. (A) KEGG enrichment of MRM-identified DEPs between AMS4k and AMS1k with p-values less than 0.05. Glycolysis/gluconeogenesis, carbon metabolism, and HIF-1 signaling pathways were involved in the development of AMS. (B) KEGG enrichment of the DEPs between the nAMS4k and nAMS1k groups with p-values less than 0.05. The hematopoietic cell lineage was mainly involved in individuals without AMS exposed to high altitude.

**Figure S3:**
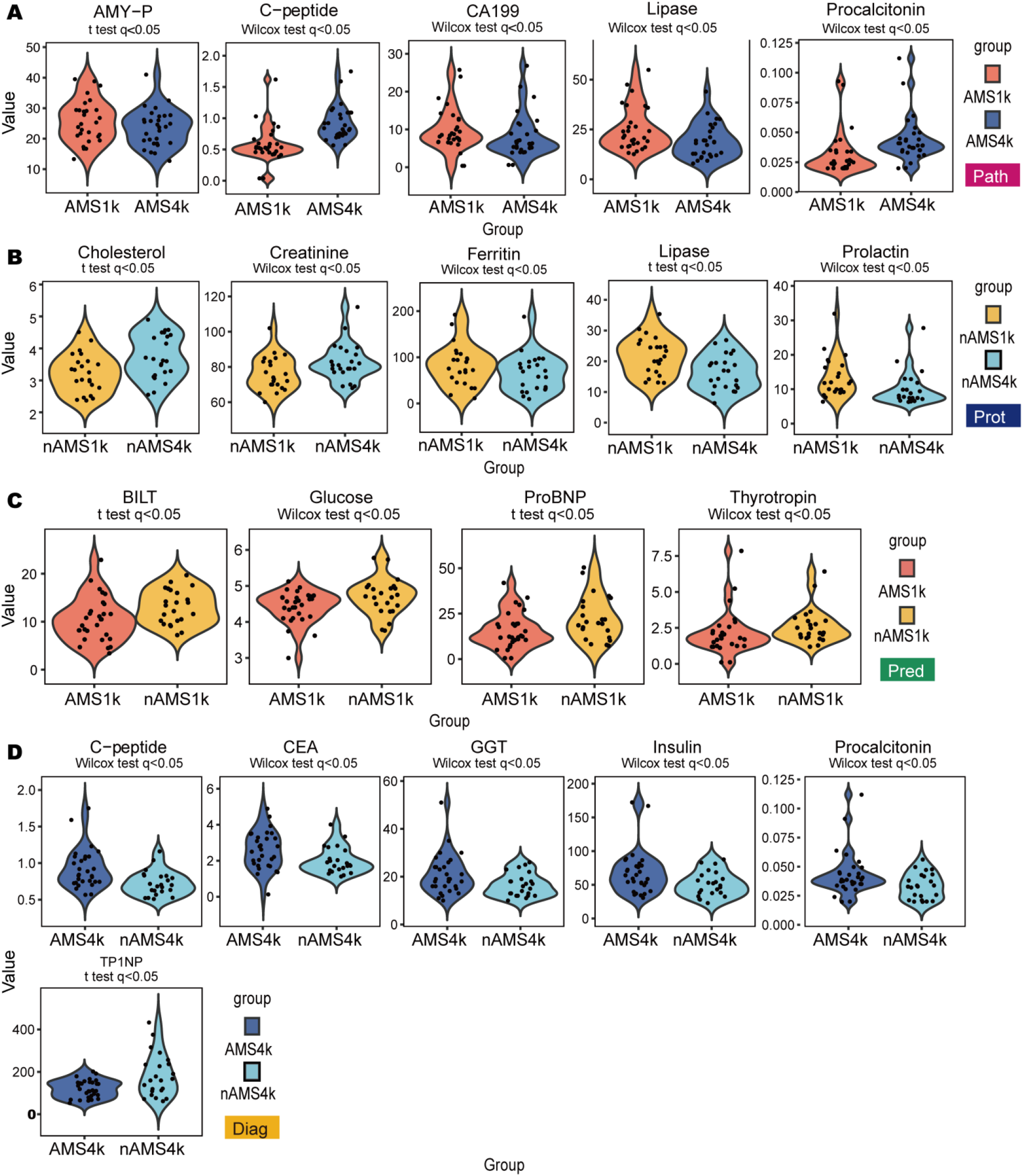
Violin plots of key clinical indexes with statistical significance. (A) Violin plots of five selected clinical indexes showing differences between AMS1k (red violin) and AMS4k (blue violin) (pathogenesis, pink box). (B) Violin plots of five selected clinical indexes showing differences between nAMS4k (light green violin) and nAMS1k (light yellow violin) (protection, dark blue box). (C) Violin plots of clinical indexes showing differences between AMS1k (blue violin) and nAMS1k (light yellow violin) (prediction, dark green box). (D) Violin plots of six clinical indexes showing differences between the AMS4k (red violin) and nAMS4k (light green violin) groups (diagnosis, yellow box). The comparisons between two groups were assessed by paired or unpaired t-tests and Wilcoxon tests (top of the violin plot) when appropriate.

**Figure S4:**
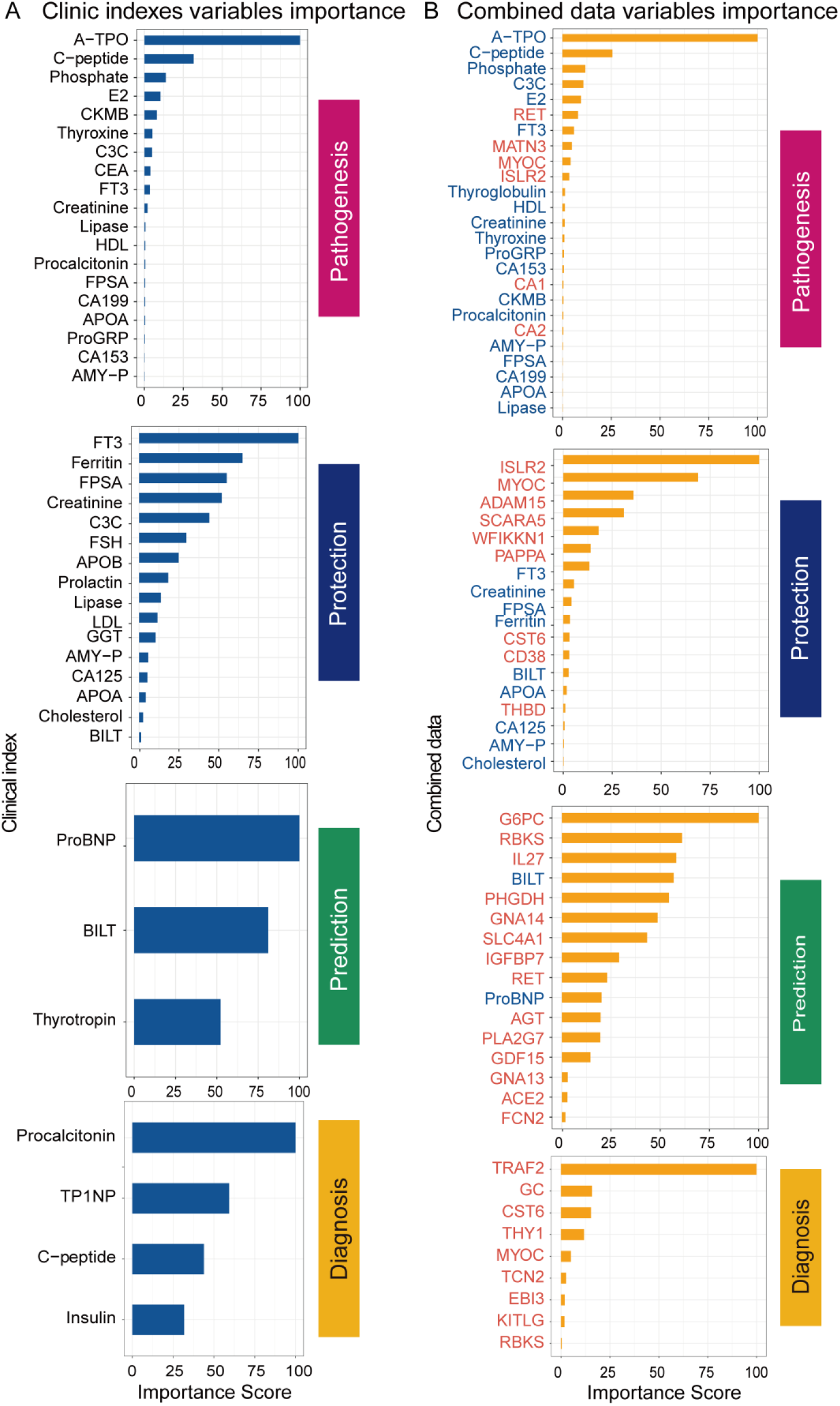
Bar plots of important features in the machine learning models. (A) Bar plots of the importance of the clinical indexes (blue bars) in the pathogenesis (pink box), protection (dark blue box), prediction (dark green box), and diagnosis (yellow box) models. (B) Bar plots of the importance of the combined data (yellow bars) in the pathogenesis (pink box), protection (dark blue box), prediction (dark green box), and diagnosis (yellow box) models. Proteins (orange fonts) accounted for a relatively higher proportion of the important features compared with clinical indexes (blue fonts) in the four comparison models established using the combined data.

**Figure S5:**
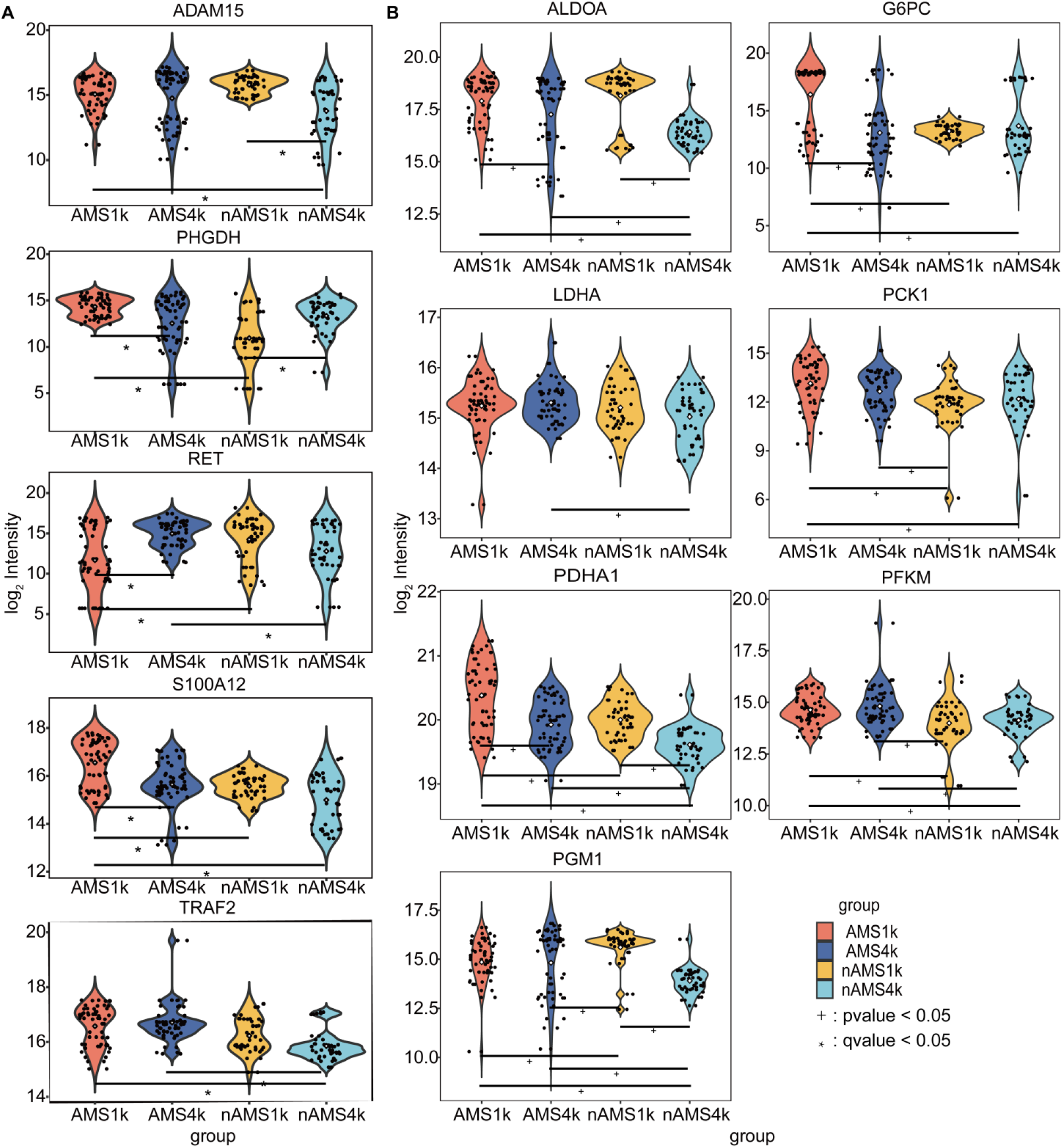
Violin plots of the comparison analysis of key proteins involved in carbohydrate metabolism. (A) Violin plot of ADAM15, S100A12, PHGDH, RET and TRAF2 proteins in the AMS1k (red violin) and AMS4k (blue violin), nAMS1k (light yellow violin) and nAMS4k (light green violin) groups. (B) Violin plot of 7 proteins related to carbohydrate metabolism. The proteins are presented in the following order: AMS1k (blue violin), AMS4k (red violin), nAMS1k (light yellow violin) and nAMS4k (light green violin). The proteins were labeled with * (q-value < 0.05) and + (p-value < 0.05). Each protein related to carbohydrate metabolism was significantly changed in at least one comparison.

**Figure S6:**
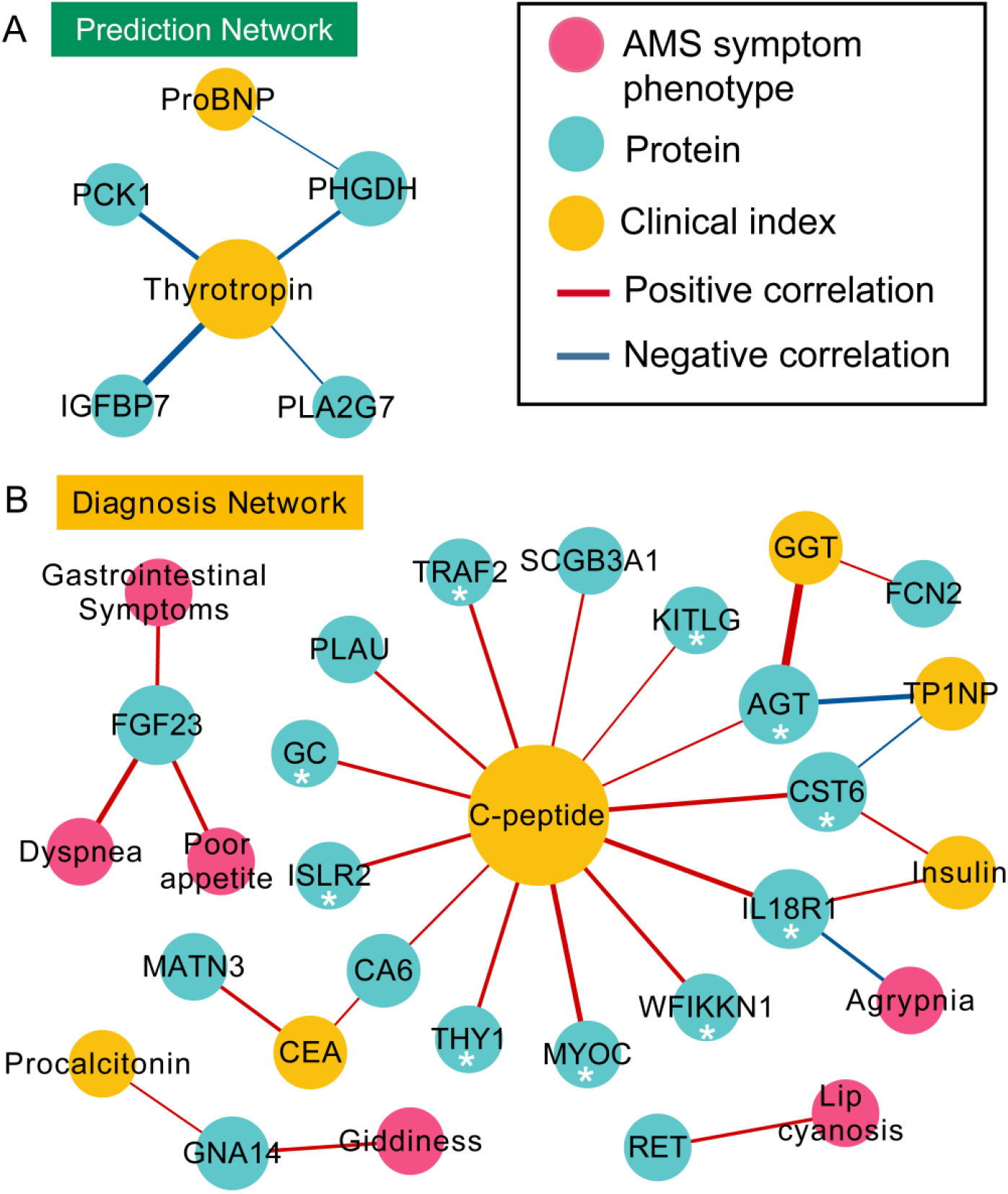
Network visualization of MRM-identified DEPs, clinical indexes and AMS symptom phenotypes in the prediction and diagnosis comparisons. (A) The network of AMS symptom phenotypes (pink dots), DEPs (blue dots), and clinical indexes (yellow dots) were connected based on the Spearman correlation coefficients and the statistical significance between AMS1k and nAMSk (prediction, dark green box). (B) The network of AMS symptom phenotypes (pink dots), DEPs (blue dots), and clinical indexes (yellow dots) were connected based on the Spearman correlation coefficients and the statistical significance between AMS4k and nAMS4k (diagnosis, yellow box). The edges showing a positive correlation (red line) and a negative correlation (blue line) with q-values less than 0.05 are shown. The thickness of the edges corresponds to the absolute value of the correlation coefficients. The size of the node is proportional to the number of edges from the node. C-peptide showed positive correlation with proteins that were selected by XGBoost model or differential analysis and labeled with *.

**Table S1: List of AMS symptom phenotypes and clinical indexes. AMS symptom phenotype sheet:** Twenty AMS-related symptom phenotypes were assessed. The gastrointestinal symptom consists of poor appetite, vomiting, and nausea. The LLS was evaluated based on headache, gastrointestinal symptoms, dizziness, and fatigue. **Clinical indexes sheet:** Overall, 65 clinical indexes were detected.

**Table S2: List of 1069 proteins in 12 Olink panels**. A total of 1069 proteins in 12 Olink panels and the detectable rate of each panel are shown.

**Table S3: Statistical results for differentially expressed proteins verified by PEA and MRM. PEA DEP sheet:** The statistical results, such as the statistical method, log2FC and q-value, of DEPs identified by PEA are summarized. The upregulated (red fonts, n = 40) and downregulated (blue fonts, n = 7) proteins identified at the discovery stage are highlighted. **Pathogenesis sheet**: The statistical results of DEPs identified by PEA are listed. The proteins with the same regulation trends as that obtained for PEA-identified DEPs between AMS4k and AMS1k are highlighted (green backgrounds, n = 23). Among these proteins, proteins with q-values less than 0.05 are labeled (red fonts, n = 4). **Protection sheet:** The statistical results of DEPs identified by PEA are summarized. The DEPs between nAMS4k and nAMS1k are highlighted (green backgrounds, n = 29), and proteins showing opposite regulation trends between the AMS4k and AMS1k groups based on PEA-identified DEPs and the proteins validated by MRM are marked (red fonts, n = 5). **Prediction sheet:** The statistical results of DEPs identified by PEA are summarized. The DEPs between AMS1k and nAMS1k are highlighted (green backgrounds, n = 23). The proteins with absolute values of log2 FC larger than 0.5 and the same regulation trends as those obtained for the proteins in the comparison between the AMS4k and AMS1k groups are marked (red fonts, n = 6). **Diagnosis sheet:** The statistical results of DEPs identified by PEA are summarized. The proteins showing differential expression between AMS4k and nAMS4k but not between AMS1k and nAMS1k are highlighted (green backgrounds, n = 28). Among these proteins, proteins with absolute values of log2 FC larger than 0.5 are marked (red fonts, n = 10).

**Table S4: GOBP and KEGG enrichment results**. GOBP and KEGG enrichment results of the two clusters of proteins identified by PEA between the AMS4k and AMS1k groups and the KEGG enrichment results of the proteins validated by MRM between the AMS4k and AMS1k groups and the nAMS4k and nAMS1k groups.

**Table S5: Statistical analysis results of 65 clinical indexes. Pathogenesis sheet:** Statistical analysis results of clinical indexes between AMS4k and AMS1k. Overall, 18 indexes (green background) are also differential indexes (q-values < 0.05) between the nAMS4k and nAMS1k groups. Eight indexes (orange background) are not differential indexes between the nAMS4k and nAMS1k groups. **Prediction sheet:** Statistical analysis results between AMS1k and nAMS1k. Four indexes (orange background) are differential indexes (p-values < 0.05). **Diagnosis sheet:** Statistical analysis results between AMS4k and nAMS4k. Six indexes (orange background) are differential indexes (p-values < 0.05).

**Table S6: Spearman correlations among 22 AMS symptom phenotypes, differential clinical indexes, and DEPs validated by MRM**. The features, Spearman coefficients and p-values are listed in the pathogenesis, prediction, and diagnosis sheets, respectively. The correlations in the pathogenesis comparison with a p-value less than 0.05 are shown. The correlations in the prediction and diagnosis comparisons with q-values less than 0.05 are shown.

